# Diagnosis and clinical description of leptospirosis cases identified through hospital-based active surveillance in Puerto Rico, 2019-2021

**DOI:** 10.1101/2025.01.31.25321493

**Authors:** Claudia Munoz-Zanzi, Melissa Mueller, Natalia Coriano-Díaz, Ralph Rivera-Gutierrez, Christian Rivera-Cátala, Rachel Millán-Rodríguez, Rosa Cantres, Natalie M. Piñeiro-Falcón, Daiana M. Ríos-Núñez, Josean G. Maldonado-Alfonzo, Renee Galloway, Luis J. Santiago-Ramos, Amy Kircher, Deana Hallman, Lemuel Martinez, Robin Russel-Orama, Ilana J. Schafer

**Affiliations:** Division of Environmental Health Sciences, School of Public Health, University of Minnesota, Minneapolis, MN, USA; University of Puerto Rico, Medical Sciences Campus, Graduate School of Public Health, San Juan, Puerto Rico; Bacterial Special Pathogens Branch, Division of High-Consequence Pathogens and Pathology, Centers for Disease Control and Prevention, Atlanta, GA, USA; Strategic Partnerships and Research Collaborative, University of Minnesota, Minneapolis, MN, USA; Ryder Memorial Hospital, Humacao, Puerto Rico; Doctors’ Center Hospital, Manati, Puerto Rico; Castañer General Hospital, Lares, Puerto Rico

**Author notes:** Correspondence should be addressed to Claudia Munoz-Zanzi; Division of Environmental Health Sciences, School of Public Health, University of Minnesota, 220 Delaware Street, Minneapolis, MN 55454.

**Keywords:** Leptospirosis, diagnosis, acute febrile illness, active surveillance, clinical symptoms, Puerto Rico

## Abstract

**Introduction:** Leptospirosis is endemic to Puerto Rico, affecting people through seasonal increases and outbreaks. This report describes the diagnostic approach and clinical characteristics of leptospirosis cases identified through active undifferentiated acute febrile illness (UAFI) surveillance in Puerto Rico.

**Materials and Methods:** From 2019-2021, active surveillance was conducted in four emergency departments (ED), with standardized recruitment of patients aged ≥5 years, presenting with fever or a history of fever of unknown origin within the past two weeks. Additional cases were recruited from referrals of patients with positive leptospirosis diagnostic tests routinely ordered by clinicians. All enrolled patients were subject to a standardized diagnostic algorithm using PCR and serology to identify probable and confirmed leptospirosis cases. Patient data was collected from electronic medical records and interviews.

**Results:** Leptospirosis was diagnosed in 4% of 406 ED-enrolled UAFI patients, eight confirmed and eight probable cases. Referrals resulted in identification of 12 routinely detected cases (five confirmed and seven probable). Of the 28 cases, 71% were ≤7 days-post-onset, 71% were male, average age was 43 years old, and the most common presentations included lower back pain (82%), headache (75%), fatigue (71%), vomiting (71%), myalgia (71%), and calf pain (64%). Common clinical laboratory findings were elevated AST (78%), hyperglycemia (74%), thrombocytopenia (48%), proteinuria (62%), and hematuria (62%). Classification of four isolates identified *L. interrogans* serovar Icterohaemorrhagiae/Copenhageni (n=2), *L. borgpetersenii* serovar Ballum/Guangdong, and *L. kirschneri* (no serovar match). All but one of the leptospirosis cases met the current Council of State and Territorial Epidemiologists’ definition for clinical compatibility with leptospirosis. Hemorrhagic symptoms and acute kidney injury with jaundice were associated with being a leptospirosis case.

**Conclusions:** These findings underscore the role of leptospirosis among febrile illness in Puerto Rico, the need for increased awareness to optimize diagnosis and treatment, and further research to improve prevention and intervention strategies.

## 1. Introduction

Leptospirosis is a zoonotic disease of worldwide distribution caused by pathogenic bacteria of the genus *Leptospira*. Animal reservoirs include wild and domestic animals which shed the bacteria in their urine and contaminate soil and water. In 2015, a World Health Organization (WHO) task force estimated at least one million clinical cases and 58,900 deaths annually worldwide, corresponding to 2.9 million disability adjusted life years lost annually [1]. Leptospirosis is present at varying levels of endemicity in rural and urban settings and different biomes, with the greatest burden in humid tropical and subtropical areas [1, 2]. Occupational and recreational activities that put people in contact with infected animals or a contaminated environment are traditional risk factors [3]. When there are major environmental disruptions, such as hurricanes, heavy rainfall, and flooding, cases can increase, which can lead to outbreaks of varying magnitude [4, 5]. After infection, clinical presentation ranges from a mild and self-limiting flu-like illness to progression to severe systemic illness in some patients, which can include kidney failure, liver damage, and respiratory difficulty, among other significant effects. Early antibiotic treatment can improve outcomes and reduce illness duration. In severe cases, lack of appropriate treatment and access to intensive care leads to higher fatality rates [6]. Puerto Rico, located in the Western Caribbean, has favorable environmental conditions for leptospirosis transmission, including warm temperatures, heavy rainfall, and natural disasters such as hurricanes that result in flooding, as well as marginalized localities with inadequate infrastructure. Since the initial reports of leptospirosis in Puerto Rico dating back to 1918 [7–10] cases have remained frequent [11, 12]. Notably, Puerto Rico has consistently reported the highest number of cases compared to all other U.S. states and jurisdictions since leptospirosis was re-instated as a nationally notifiable disease in 2014 [13]. Leptospirosis has remained a mandatory reportable disease in Puerto Rico –even when the Council of State and Territorial Epidemiologists (CSTE) temporarily removed leptospirosis from the list of nationally notifiable conditions between 1995 and 2013. A study analyzing 570 confirmed cases reported to the Puerto Rico Department of Health (PRDH) between 1996-2014 estimated the highest annual incidence as 2.11/100,000 in 2007 [14]. Surveillance data show the endemicity of leptospirosis in Puerto Rico, and this is further demonstrated in investigations using samples from dengue suspected but test-negative patients, which revealed that 5% to 9% of the samples were leptospirosis positive [15, 16]. However, because underdiagnosis is common due to clinical similarity with other common infections and the need for specialized laboratory testing, reported cases likely represent an underestimation, and the true burden of the disease is unknown. An initial and critical step in improving timely and accurate detection is to identify patients with suspected infections who need leptospirosis laboratory testing [17]. Periodic outbreaks have been reported in Puerto Rico after heavy rain and floods, including post-hurricane Hortense in 1996 [11], Irma and Maria in 2017 [18], and Fiona in 2022 [19, 20]. Following the particularly devastating hurricanes Irma and Maria, the United States Centers for Disease Control and Prevention (CDC) facilitated the implementation of active hospital-based surveillance for leptospirosis to measure the burden among patients with undifferentiated acute febrile illness (UAFI) and to gain knowledge on the epidemiology, strain diversity, and exposure risk. The objective of this report is to describe the diagnosis, clinical presentation, clinical management, and outcomes of leptospirosis cases identified through active febrile illness surveillance at four hospitals in Puerto Rico.

## 2. Materials and Methods

The approach for the implementation of active hospital-based surveillance was to identify patients presenting at selected hospitals’ emergency departments (ED) with UAFI and apply a standardized diagnostic algorithm for leptospirosis. To capture as many leptospirosis cases as possible, patients referred by the hospital’s laboratory as leptospirosis cases based on a positive routine test ordered by the attending physician (due to clinical suspicion of leptospirosis) were also identified for recruitment. The protocol was approved as a public health surveillance activity with non-research determination by the CDC’s National Center for Emerging and Zoonotic Infectious Diseases Human Subjects Team (# 043019IS) and the University of Minnesota Institutional Review Board (#4743).

### 2.1 Recruitment and Enrollment

Four hospitals were selected based on location, patient volume, laboratory capacity for on-site testing, and ability to allow project personnel to implement the project’s processes on-site. The hospitals were Doctors’ Center Hospital (DCH) in Manatí with 258 beds, Ryder Memorial Hospital (RMH) in Humacao with 167 beds, Castañer General Hospital (CGH) in Lares with 24 beds, and Bella Vista Hospital (BVH) in Mayagüez with 157 beds (Figure 1). The overall project enrollment period was from July 2019 through May 2021; however, enrollment time by hospital varied depending on the respective start time and COVID-19 pandemic interruptions. Enrollment periods were from July 2019 - March 2021 for DCH, from December 2019 - April 2021 for CGH, from February 2020 - March 2020 for BVH, and from March 2020 - May 2021 for RMH. All sites paused enrollment for approximately three months from mid-March 2020 through June 2020 due to the COVID-19 pandemic and the need to adjust protocols to the hospitals’ new safety, triage, and patient isolation procedures. Enrollment at BVH did not restart after the pause due to administrative reasons. Total active enrollment periods were 18 months for DCH, 13 months for CGH, 1.5 months for BVH, and 11.5 months for RMH. Eligible patients were identified and recruited at each hospital by project coordinators based on systematically searching ED triage information for patients meeting the pre-screening criteria of ≥5 years of age and either current fever (tympanic/oral temperature ≥38°C or temporal/axillary ≥37.5°C) and/or self-reported history of fever any time in the past two weeks. This process was carried out five days per week. Recruitment shifts varied between morning (7 a.m. to 3 p.m.) and/or evening (3 p.m. to 11 p.m.) and from one to two shifts per day, depending on the number of project coordinators assigned to each site.

**FIGURE 1:**
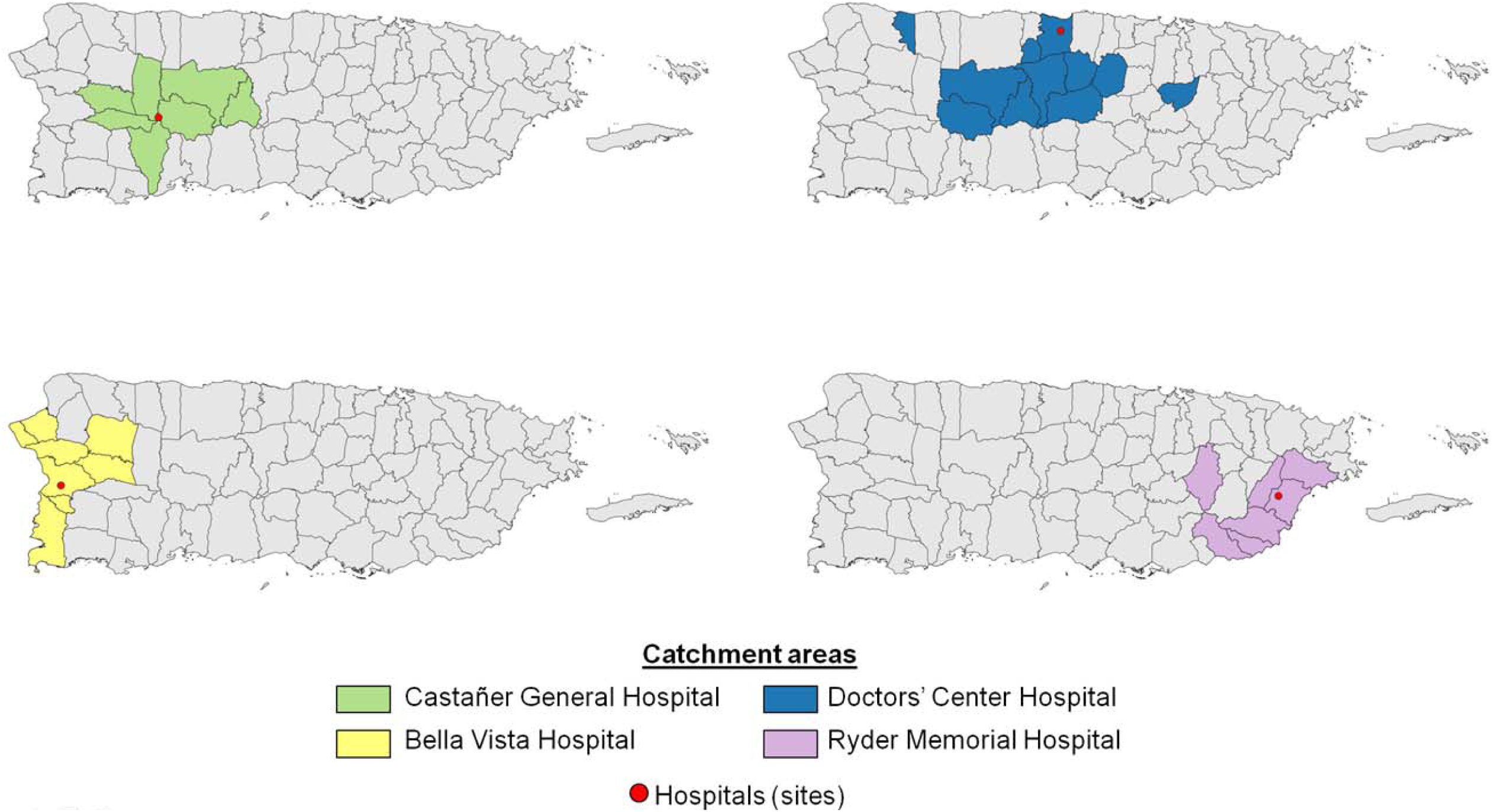
Four hospitals conducting active surveillance for leptospirosis through emergency room visits and their corresponding catchment areas, 2019-2021, Puerto Rico.

Project coordinators located the patients meeting pre-screening criteria on the ED floor and completed full enrollment screening. Patients who met the following inclusion criteria, and did not meet exclusion criteria, were invited to enroll: i) ≥5 years old, ii) current fever or self-reported history of fever in the past two weeks, and iii) illness onset within the past two weeks. Exclusion criteria included a likely non-infectious and/or predisposing condition that could cause fever such as cancer, recent surgery, recent trauma/wounds, or an abscess; a laboratory confirmed diagnosis for another infection that could reasonably cause the current illness; previous enrollment in the project for the same illness; and inability to provide consent (minors, mentally impaired individuals, unconscious/too sick to understand consent form) and not accompanied by a parent or legal guardian. Those identified as eligible were enrolled after written informed consent and/or assent was provided by the patient, and/or a proxy or legal guardian depending on the age and ability of the patient to provide consent/assent. These patients are referred to as ED-enrolled patients.

A separate process was established to identify and enroll patients throughout the hospital who tested positive for leptospirosis on a routine diagnostic test ordered by the attending physician independent of the active surveillance enrollment process in the ED. Project coordinators were notified by the hospital’s laboratory of any patient who had a leptospirosis IgM positive or borderline test or a PCR positive test performed at the hospital or a reference laboratory. These referrals included patients seen at the hospital ED since the beginning of the project and at any time during and outside project recruitment hours. These enrolled patients are referred to as laboratory-positive referrals. All laboratory-positive referrals were invited to enroll after informed consent/assent was obtained.

### 2.2. Specimen collection

Leptospirosis cases were identified using a combination of PCR, culture, and serologic diagnostic tests applied in a standardized algorithm (Figure 2). If the patient was in the first week of illness (<7 days post-onset or DPO), specimens collected included serum for anti-*Leptospira* spp. antibody testing and whole blood for *Leptospira* spp. PCR testing. If the patient was in the second week of illness (7-14 DPO), specimens collected were serum for antibody testing and urine for PCR testing. Collection of a convalescent serum sample, 10-14 days or later after the acute sample, was attempted in all enrolled patients that were PCR-negative. Approaches varied by site and included asking patients to return to the hospital and/or conducting home visits by a project-supported nurse. The protocol also included the collection of a serum sample at the time of discharge for those admitted to the hospital for longer than 24 hours. Blood or urine samples from enrolled patients meeting specific clinical criteria, who had not been given antibiotics before the time of sample collection, and for whom culture inoculation could be done within one hour of sample collection, were also used for *Leptospira* spp. culture. Clinical criteria for culture for the first 5 months of enrollment starting in July 2019 included clinician-suspected leptospirosis (as entered in the electronic medical record or EMR), and/or any of the following: jaundice and/or elevated bilirubin, conjunctival suffusion, clinically suspected renal insufficiency or failure (e.g. anuria, oliguria, azotemia/elevated creatinine), clinically suspected aseptic meningitis, hemorrhage (e.g. pulmonary, hematuria, hematemesis, blood in stool, petechiae/ecchymosis), acutely elevated liver enzymes/transaminitis, calf pain, rash. In December 2019, the list was expanded to include conjunctivitis, gastrointestinal symptoms (diarrhea, vomiting, nausea), meningitis (vs. aseptic meningitis), thrombocytopenia, and reports of headache and myalgia together.

**FIGURE 2:**
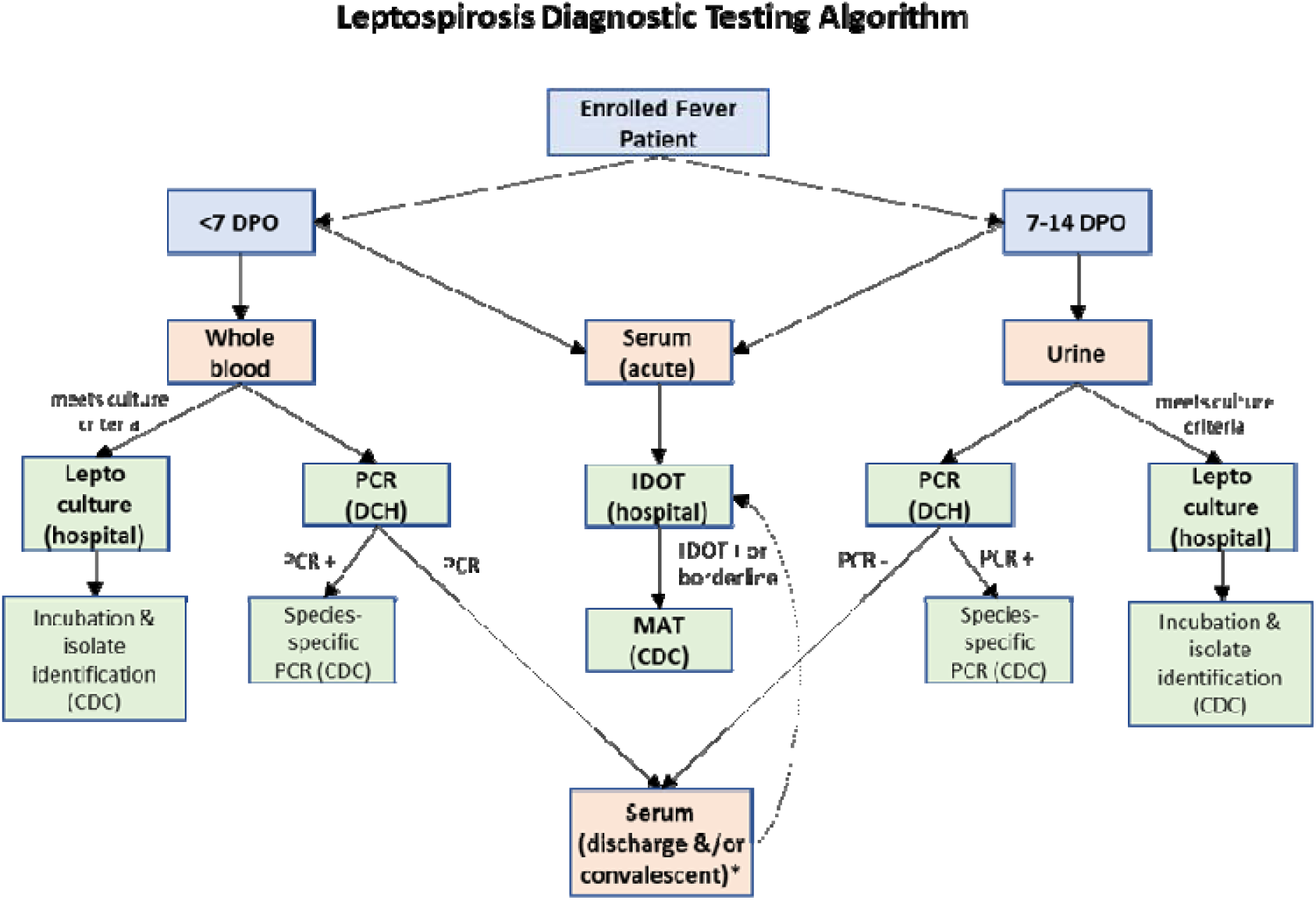
Specimen collection and diagnostic testing algorithm for leptospirosis applied to patients meeting acute febrile illness enrollement criteria at four hospitals in Puerto Rico, 2019-2021. DPO: Days post onset. DCH: Doctor’s Center Hospital clinical laboratory in Puerto Rico performing the leptospirosis PCR test. CDC: Centers for Disease Control and Prevention, United States, performing the microagglutination (MAT) test. Outpatients had a convalescent serum sample collected 10-14 days after the acute serum sample. Inpatients hospitalized for ≥ 24 hours had a discharge serum sample collected. If the discharge sample was collected < 7 days after the acute serum sample, a convalescent sample collection was also attempted 10-14 days after acute specimen collection.

### 2.3 Leptospirosis laboratory diagnosis

An anti-*Leptospira* IgM enzyme-linked immunoassay (IgM ImmunoDOT^TM^ *Leptospira* test, GenBio, San Diego, CA) (IDOT), commercially available and FDA approved, was run on all serum samples at each site’s clinical laboratory following manufacturers’ instructions. IDOT test results were interpreted as negative, borderline, or positive. An in-house *lipL32* PCR protocol conducted at CDC until February 2021 and at Doctors’ Center Hospital thereafter adapting a similar protocol, provided positive or negative results for *Leptospira* spp. DNA [21, 22]. Blood and urine specimens for PCR testing were transported from the other three sites to DCH by project personnel on a routine schedule. The following samples were submitted to CDC for *Leptospira* microscopic agglutination test (MAT): acute, discharge, and/or convalescent serum samples with positive or borderline IDOT results and convalescent samples of any patient with an IDOT borderline or positive result in the acute sample (regardless of their IDOT result in the convalescent sample). The MAT was run as previously described using a panel of 20 reference strains [23]. Media for *Leptospira* spp. culture was obtained from USDA and consisted of two media [24, 25]. each run in duplicate. Whole blood and urine-inoculated media tubes were transported to the CDC to complete an incubation period of up to 6 months. Species identification for PCR-positive samples was done as described previously [26] using PerfeCTa Tough Mix, Low ROX (Quantabio, Beverly, MA). Isolates were classified at the species and serovar level using PFGE [27].

### 2.4 Leptospirosis case definition

Each enrolled patient was classified based on the leptospirosis laboratory test results. A patient was classified as a confirmed case when any of the following criteria were met: PCR positive on urine or blood, MAT titer ≥1:800 in a single serum specimen, ≥4-fold increase in MAT titers between acute and convalescent (or discharge) serum samples, or culture-positive results. A probable case was one with a MAT titer ≥1:200 but <1:800 and/or IgM-positive result on a serum specimen. A patient was classified as “not a case” when a convalescent serum sample collected ≥14 DPO was IgM and/or MAT negative. Patients who did not meet any of these categories were classified as “fever patients”.

### 2.5 Data collection

Patient data were collected at different time points by project coordinators using standardized forms. At the time of enrollment, site coordinators interviewed the patient or a legal representative to obtain information on demographics and living conditions, medical history, symptoms, prior treatment, and possible exposures and risk factors. Data on the ED clinical exam findings, preliminary diagnosis, disposition after the ED visit, routine diagnostic tests (complete blood count, chemistry, urinalysis), non-leptospirosis infectious disease tests, antibiotic and supportive treatments provided, and final diagnosis at discharge were extracted from the electronic medical records. For enrolled patients admitted to the hospital, clinical data were collected from the time of the ED visit as well as during hospitalization. Follow-up information was collected from patients that could be contacted approximately 10-14 days or later after the ED visit, including patient outcome, illness evolution, and supplemental follow-up care and treatment obtained. Data were entered into a project database housed in the Research Electronic Data Capture (REDCap) platform [28].

### 2.6 Data analysis

The analysis for this report aimed to describe the socio-demographic features, illness presentation, clinical management, and outcome of confirmed and probable cases, including notable differences between those identified through ED enrollment or the laboratory-positive referral process. Descriptive data summaries included counts, proportions and 95% confidence intervals and, when sample size allowed, comparisons by case classification and by enrollment method were done using chi-square tests. We assessed the proportion of cases that met the 2013 Council of State and Territorial Epidemiologists (CSTE) clinical case criteria compatible with leptospirosis [29] which include a history of fever in the past two weeks with either i) at least two of the following clinical findings: myalgia, headache, jaundice, conjunctival suffusion without purulent discharge, or rash (i.e. maculopapular or petechial) or ii) at least one of aseptic meningitis, GI symptoms (e.g., abdominal pain, nausea, vomiting, diarrhea), pulmonary complications (e.g., cough, breathlessness, hemoptysis), cardiac arrhythmias, ECG abnormalities, renal insufficiency (e.g., anuria, oliguria), hemorrhage (e.g., intestinal, pulmonary, hematuria, hematemesis), or jaundice with acute renal failure. For reporting purposes, the first criterion was referred to as CSTE AFI and the second one as CSTE Severe. The performance of these criteria to identify leptospirosis cases for surveillance based on clinical signs at the ED visit was evaluated by estimating the sensitivity (SE) and specificity (SP) and their 95% exact confidence intervals and predictive values positive and negative. To identify sets of symptoms that were associated with being a leptospirosis case (confirmed or probable), the prevalence of leptospirosis was estimated among those with and without the symptoms to estimate a prevalence ratio (PR) and its 95% confidence interval. For these analyses, new variables were created to combine the symptom information obtained from the patient interview and the information extracted from medical records at the time of the ED visit, as appropriate. If a given symptom was present in either source, the symptom was considered present. The R package epiR was used for statistical analyses [30] within the RStudio platform [31].

## 3. Results

### 3.1 Enrollment and leptospirosis diagnosis

Project coordinators identified a total of 1,689 patients as meeting the pre-screening acute febrile illness criteria based on ED triage records. Of those, 1,158 (68.6%) patients, or their proxies, were found by the recruiters and were further screened for enrollment eligibility. The final number of ED-enrolled patients who met eligibility criteria and consented to participate was 406 (54.6% of the 744 eligible). A total of 14 laboratory-positive patients, all initially seen at the ED, were referred to the coordinators and 13 (92.9%) were enrolled (Table 1). The application of the laboratory testing algorithm to the 406 ED-enrolled patients identified 16 (3.9%) leptospirosis cases – eight of which were confirmed and eight probable cases. Among the rest, 79 (19.5%) were classified as not a leptospirosis case based on a seronegative convalescent sample, and 311 (76.6%) were classified as fever patients, indicating they did not meet any other case definition. Of the 13 laboratory-positive referral patients, five (38.6%) were classified as confirmed cases and seven (53.9%) as probable cases. One patient (7.7%), referred because of an initial IgM borderline result, was classified as not a leptospirosis case after a MAT negative result on a convalescent sample. Overall, there were 28 leptospirosis cases (13 confirmed and 15 probable) identified by the entire active surveillance investigation.

**TABLE 1:**
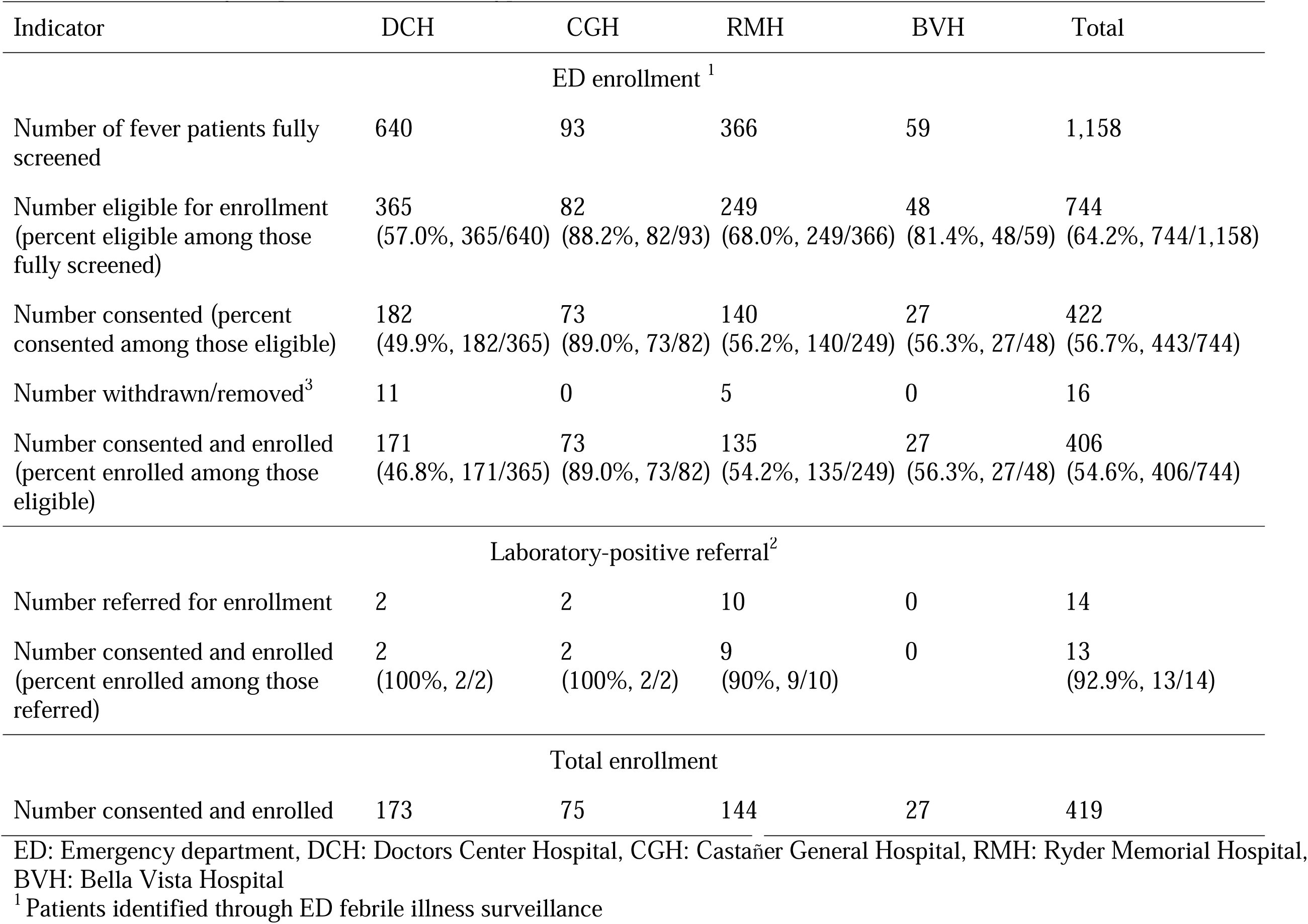
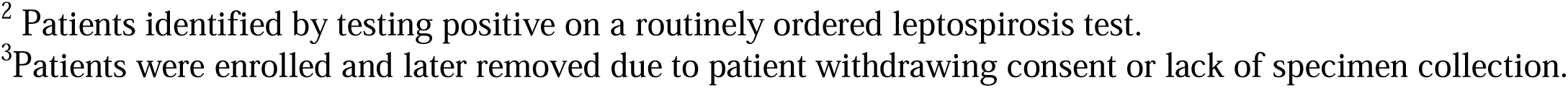
Summary of screening and enrollment of emergency room patients in a leptospirosis active surveillance project in Puerto Rico, broken down by hospital and enrollment type, 2019-2021.

| Indicator | DCH | CGH | RMH | BVH | Total |
| --- | --- | --- | --- | --- | --- |
| ED enrollment <sup>1</sup> |  |  |  |  |  |
| Number of fever patients fully screened | 640 | 93 | 366 | 59 | 1,158 |
| Number eligible for enrollment<br>(percent eligible among those fully screened) | 365<br>(57.0%, 365/640) | 82<br>(88.2%, 82/93) | 249<br>(68.0%, 249/366) | 48<br>(81.4%, 48/59) | 744<br>(64.2%, 744/1,158) |
| Number consented (percent consented among those eligible) | 182<br>(49.9%, 182/365) | 73<br>(89.0%, 73/82) | 140<br>(56.2%, 140/249) | 27<br>(56.3%, 27/48) | 422<br>(56.7%, 443/744) |
| Number withdrawn/removed<sup>3</sup> | 11 | 0 | 5 | 0 | 16 |
| Number consented and enrolled<br>(percent enrolled among those eligible) | 171<br>(46.8%, 171/365) | 73<br>(89.0%, 73/82) | 135<br>(54.2%, 135/249) | 27<br>(56.3%, 27/48) | 406<br>(54.6%, 406/744) |
| Laboratory-positive referral<sup>2</sup> |  |  |  |  |  |
| Number referred for enrollment | 2 | 2 | 10 | 0 | 14 |
| Number consented and enrolled<br>(percent enrolled among those referred) | 2<br>(100%, 2/2) | 2<br>(100%, 2/2) | 9<br>(90%, 9/10) | 0 | 13<br>(92.9%, 13/14) |
| Total enrollment |  |  |  |  |  |
| Number consented and enrolled | 173 | 75 | 144 | 27 | 419 |

ED: Emergency department, DCH: Doctors Center Hospital, CGH: Castañer General Hospital, RMH: Ryder Memorial Hospital, BVH: Bella Vista Hospital
<sup>1</sup> Patients identified through ED febrile illness surveillance
<sup>2</sup> Patients identified by testing positive on a routinely ordered leptospirosis test.
<sup>3</sup> Patients were enrolled and later removed due to patient withdrawing consent or lack of specimen collection.

Among the 13 confirmed cases, four (30.8%) were PCR positive only, four (30.8%) were both PCR positive and had a ≥4-fold MAT titer increase, one (7.6%) was PCR positive and had a MAT titer of 1:200-1:400, one (7.6%) was confirmed through ≥4-fold MAT titer increase alone, and three (23.1%) had a single MAT titer ≥1:800. All PCR positive results were on blood samples. Nine of the 13 (69.2%) confirmed cases were IDOT IgM positive, all of which were eventually MAT positive. All 15 probable cases (100%) were identified through IDOT IgM, with two of them having a MAT titer of 1:100. All eight (100%) of the ED-enrolled confirmed cases were PCR positive, while one of the five (20%) laboratory-positive referral confirmed cases were PCR positive (Table 2).

**TABLE 2:**
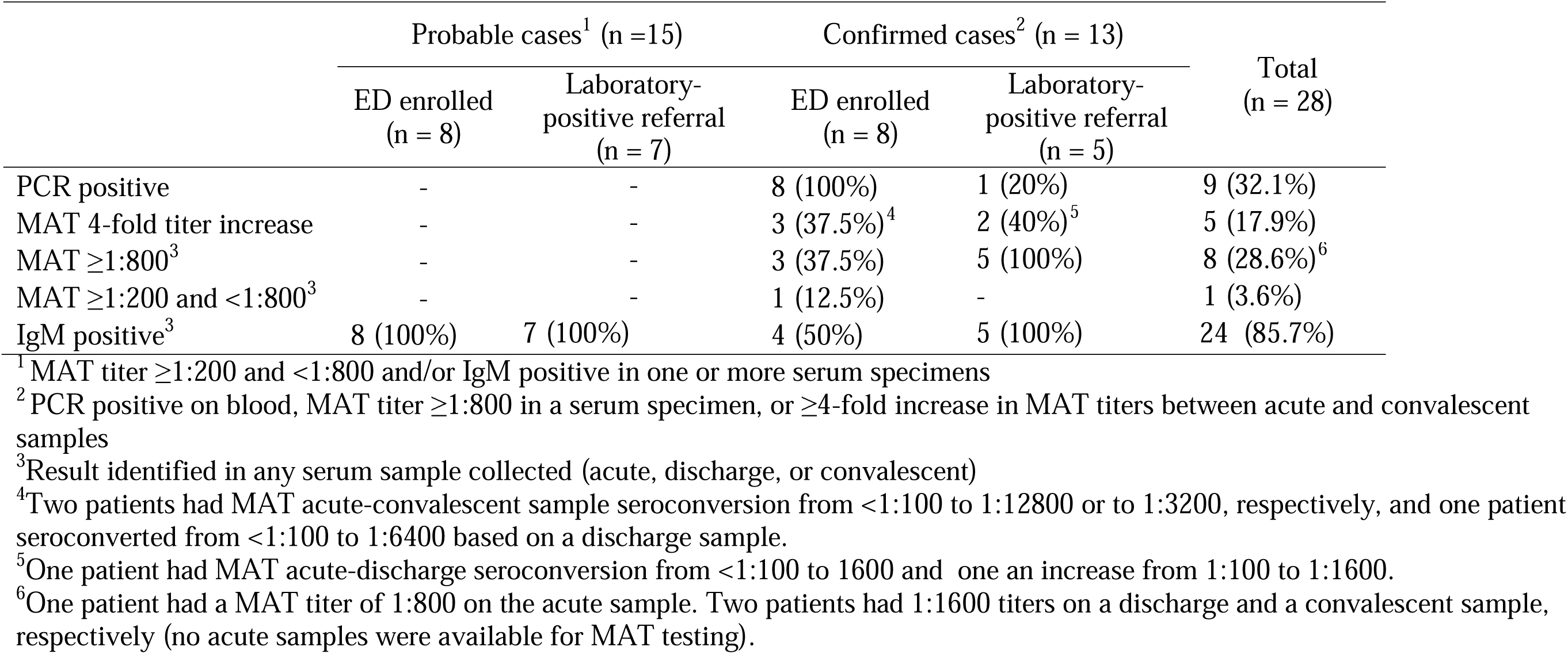
Summary of laboratory diagnostic results of leptospirosis cases identified through active hospital-based surveillance in Puerto Rico, 2019-2021.

Confirmed cases who had ≥4-fold MAT titer increase or had MAT titers ≥1:200 had highest-reacting titers to serogroup Icterohaemorrhagiae (4 cases, highest titers 1:400 to 1:12,800), serogroup Canicola (2 cases, highest titers 1:800 and 1:6,400), serogroup Ballum (1 case, highest titer 1:1,600), serogroup Bataviae (1 case, highest titer 1:1,600), and serogroups Icterohaemorrhagiae and Canicola (1 case, highest titer 1:3,200 to both).

Species classification by PCR was attempted on five of the nine PCR-positive samples. Three were *L. interrogans*, one was *L. borgpetersenii*, and one was *L. kirschneri*. Specimens from 68 enrolled patients were cultured and four were positive. For two of these cases, the isolates were classified as *L. interrogans* serovar Icterohaemorrhagiae/Copenhageni and both were PCR positive, IDOT IgM positive, and seroconverted on MAT. The highest MAT titers were to serogroups Icterohaemorrhagiae and Canicola equally for one of these culture-positive patients, and to serogroup Canicola for the second. The other two isolates were classified as *L. borgpetersenii* serovar Ballum/Guangdong and *L. kirschneri* (no serovar match by PFGE), respectively, and both cases were PCR positive and IDOT IgM negative (MAT testing was not done).

### 3.2 Epidemiological characteristics

Overall, the average age of the cases was 43.1 years and 71.4% (20/28) were male. All 28 leptospirosis cases considered themselves Hispanic and Puerto Ricans and had permanent residency in Puerto Rico. Among 11 cases who reported income, all had an income < USD $40,000/year, and for 63.6% (7/11) it was under ≤ USD $20,000/year. Most cases used piped or bottled water and only one case (confirmed case) reported use of water sourced from an open well for drinking and bathing. Over half (15/27, 53.5%) reported the occurrence of heavy rain at their home or work within the past 30 days, and 17.9% (5/28) reported flooding. Two cases, one probable and one confirmed case, reported contact with flood water. Overall, reported activities that resulted in contact with water or mud/soil included yard work, laundry or washing dishes, taking care of pets (dogs/cats) or farm animals, playing sports in the yard or park, construction at home, and cleaning after an earthquake or flood. Seven cases (7/28, 25%) had open wounds or cuts in the previous 30 days, six of whom were confirmed cases. The most common animal exposure or contact was the presence of rodents at home or work (19/26, 73.1%), particularly for confirmed cases (12/13, 92.3%), followed by contact with dogs (18/28, 64.3%) (Table S1). One confirmed case-patient identified through active surveillance reported a previous history of leptospirosis and one probable case reported travel outside Puerto Rico but within the U.S. in the past 30 days.

### 3.3 Clinical presentation at the ED visit

Most cases (20/28, 71.4%) sought care during the first week of illness (DPO ≤ 7) at a mean of 6.3 days (range: 0 – 27). A greater proportion of ED- enrolled cases (15/16, 93.8%) had a DPO ≤ 7 days than laboratory-positive referral cases (5/12, 41.7%) (p = 0.07) at the time of interview. Thirty-one percent (5/16) of ED-enrolled cases sought care for the same illness prior to the ED visit; this proportion was greater for laboratory-positive referral cases (7/12, 58.3%). Similarly, a greater proportion of laboratory-positive referral cases (4/12, 33.3%) were prescribed antibiotics prior to the ED visit compared with ED-enrolled cases (2/16, 12.5%).

The most common symptoms at the time of presentation among the 13 confirmed cases were lower back pain (92%), myalgia (92%), fatigue or weakness (92%), vomiting or nausea (85%), headache (77%), calf pain (77%), and arthralgia (77%). Among the 15 probable cases, the most common symptoms were headache (73%), lower back pain (73%), stiff neck (60%), and vomiting/nausea (60%) (Table 4). When comparing the 13 confirmed and 15 probable cases, regardless of enrollment method, fatigue/weakness (92% vs 53%, p = 0.038), myalgia (92% vs 53%, p = 0.038), diarrhea (62% vs 27%, p = 0.125), and arthralgia (77% vs 47%, p = 0.137) were more common among confirmed than probable cases (Table 3). There were notable but not statistically significant differences in symptoms between the five laboratory-positive referral confirmed cases and the eight ED-enrolled confirmed cases. More laboratory-positive referral confirmed cases than ED-enrolled confirmed cases had abdominal pain (100% vs 38%, p-value = 0.075), red eyes (80% vs 25%, p = 0.103), and hematuria (60% vs 13%, p = 0.217). Conversely, there were more ED-enrolled confirmed cases than laboratory-positive referral confirmed cases with current fever (75% vs 20%, p = 0.103). There were more laboratory-positive referral probable cases than ED-enrolled probable cases with fatigue/weakness (86% vs 25%, p = 0.041), abdominal pain (71% vs 25%, p = 0.132), light sensitivity (71% vs 25%, p = 0.132), dyspnea/shortness of breath (43% vs 0%, p = 0.077), and oliguria/anuria (43% vs 0%, p = 0.077).

**TABLE 3:** Illness presentation and healthcare-seeking behavior at the time of the ED visit among leptospirosis cases identified through hospital-based active surveillance in Puerto Rico, 2019-2021.

|  | Leptospirosis cases |  |  |  |  |  |  |
| --- | --- | --- | --- | --- | --- | --- | --- |
|  | Probable |  |  | Confirmed |  |  | Total leptospirosis cases (n = 28) |
|  | ED enrolled (n = 8) | Laboratory-positive referral (n = 7) | Probable Total (n = 15) | ED enrolled (n = 8) | Laboratory-positive referral (n = 5) | Confirmed Total (n = 13) |  |
| DPO average (range) | 3.5 (1-7) | 14.0 (0-27) | 8.4 (0-27) | 2.4 (1-4) | 6.2 (4-12) | 3.9 (1-12) | 6.3 (0-27) |
| DPO $\leq 7$ days | 7/8 (87.5%) | 2/7 (28.6%) | 9/15 (60%) | 8/8 (100%) | 3/5 (60%) | 11/13 (84.6%) | 20/28 (71.4%) |
| Sought care prior to ED visit | 2/8 (25%) | 4/7 (57.1%) | 6/15 (40%) | 3/8 (37.5%) | 3/5 (60%) | 6/13 (46.2%) | 12/28 (42.9%) |
| Antibiotic prior to ED visit | 2/8 (25%) | 3/7 (42.9%) | 5/15 (33.3%) | 0/8 | 1/5 (20%) | 1/13 (7.7%) | 6/28 (21.4%) |
| BP systolic average (range) | 125.8 (110-176) | 124.4 (93- 153) | 125.1 (93-176) | 121.4 (101-158) | 119.6 (88-145) | 120.7 (88-158) | 123.1 (88-176) |
| BP diastolic average (range) | 77.9 (70-97) | 74.6 (52-96) | 76.3 (52-97) | 70.9 (56-81) | 76.6 (56-88) | 73.1 (56-88) | 74.8 (52-97) |
| Heart rate average (range) | 94 (73-113) | 109.3 (76-150) | 101.1 (73-150) | 94 (74-108) | 90.8 (70-124) | 92.8 (70-124) | 97.3 (70-150) |
| Respiratory rate average (range) | 17.6 (16-19) | 18.1 (17-19) | 17.9 (16-19) | 18.8 (17-22) | 20.6 (18-28) | 19.5 (17-28) | 18.6 (16-28) |
| Current fever | 2/8 (25%) | 3/7 (42.9%) | 5/15 (33.3%) | 6/8 (75%) | 1/5 (20%) | 7/13 (53.9%) | 12/28 (42.9%) |
| Chills | 0/8 | 0/7 | 0/15 | 0/8 | 1/5 (20%) | 1/13 (7.7%) | 1/28 (3.6%) |
| Headache | 6/8 (75%) | 5/7 (71.4%) | 11/15 (73.3%) | 5/8 (62.5%) | 5/5 (100%) | 10/13 (76.9%) | 21/28 (75%) |
| Fatigue/weakness | 2/8 (25%) | 6/7 (85.7%) | 8/15 (53.3%) | 7/8 (87.5%) | 5/5 (100%) | 12/13 (92.3%) | 20/28 (71.4%) |
| Disoriented/altered mental status | 3/8 (37.5%) | 4/7 (57.1%) | 7/15 (46.7%) | 3/8 (37.5%) | 2/5 (40%) | 5/13 (38.5%) | 12/28 (42.9%) |
| Myalgia | 5/8 (62.5%) | 3/7 (42.9%) | 8/15 (53.3%) | 7/8 (87.5%) | 5/5 (100%) | 12/13 (92.3%) | 20/28 (71.4%) |
| Malaise/body ache | 0/8 | 0/7 | 0/15 | 2/8 (25%) | 1/5 (20%) | 3/13 (23.1%) | 3/28 (10.7%) |
| Calf pain | 3/8 (37.5%) | 5/7 (71.4%) | 8/15 (53.3%) | 6/8 (75%) | 4/5 (80%) | 10/13 (76.9%) | 18/28 (64.3%) |
| Lower back pain | 6/8 (75%) | 5/7 (71.4%) | 11/15 (73.3%) | 7/8 (87.5%) | 5/5 (100%) | 12/13 (92.3%) | 23/28 (82.1%) |
| Arthralgia | 4/8 (50%) | 3/7 (42.9%) | 7/15 (46.7%) | 6/8 (75%) | 4/5 (80%) | 10/13 (76.9%) | 17/28 (60.7%) |
| Chest pain | 3/8 (37.5%) | 3/7 (42.9%) | 6/15 (40%) | 5/8 (62.5%) | 2/5 (40%) | 7/13 (53.9%) | 13/28 (46.4%) |
| Abdominal pain | 2/8 (25%) | 5/7 (71.4%) | 7/15 (46.7%) | 3/8 (37.5%) | 5/5 (100%) | 8/13 (61.5%) | 15/28 (53.6%) |
| Stiff neck | 4/8 (50%) | 5/7 (71.4%) | 9/15 (60%) | 3/8 (37.5%) | 2/5 (40%) | 5/13 (38.5%) | 14/28 (50%) |
| Vomiting/nausea | 4/8 (50%) | 5/7 (71.4%) | 9/15 (60%) | 6/8 (75%) | 5/5 (100%) | 11/13 (84.6%) | 20/28 (71.4%) |
| Diarrhea | 1/8 (12.5%) | 3/7 (42.9%) | 4/15 (26.7%) | 4/8 (50%) | 4/5 (80%) | 8/13 (61.5%) | 12/28 (42.9%) |
| Red eyes | 1/8 (12.5%) | 3/7 (42.9%) | 4/15 (26.7%) | 2/8 (25%) | 4/5 (80%) | 6/13 (46.2%) | 10/28 (35.7%) |
| Light sensitivity | 2/8 (25%) | 5/7 (71.4%) | 7/15 (46.7%) | 3/8 (37.5%) | 1/5 (20%) | 4/13 (30.8%) | 11/28 (39.3%) |
| Eye pain | 3/8 (37.5%) | 2/7 (28.6%) | 5/15 (33.3%) | 3/8 (37.5%) | 4/5 (80%) | 7/13 (53.9%) | 12/28 (42.9%) |
| Cough | 4/8 (50%) | 3/7 (42.9%) | 7/15 (46.7%) | 3/8 (37.5%) | 2/5 (40%) | 5/13 (38.5%) | 12/28 (42.9%) |
| Dyspnea/shortness of breath | 0/8 | 3/7 (42.9%) | 3/15 (20%) | 3/8 (37.5%) | 1/5 (20%) | 4/13 (30.8%) | 7/28 (25%) |
| Coughing blood | 0/8 | 1/7 (14.3%) | 1/15 (6.7%) | 0/8 | 0/5 | 0/13 | 1/28 (3.6%) |
| Hematuria | 0/8 | 2/7 (28.6%) | 2/15 (13.3%) | 1/8 (12.5%) | 3/5 (60%) | 4/13 (30.8%) | 6/28 (21.4%) |
| Hematemesis | 0/8 | 1/7 (14.3%) | 1/15 (6.7%) | 0/8 | 1/5 (20%) | 1/13 (7.7%) | 2/28 (7.1%) |
| Blood in stool | 0/8 | 2/7 (28.6%) | 2/15 (13.3%) | 0/8 | 0/5 | 0/13 | 2/28 (7.1%) |
| Bloody nose | 0/7 | 1/7 (14.3%) | 1/14 (7.1%) | 0/6 | 0/5 | 0/11 | 1/25 (4%) |
| Rash/petechia | 0/8 | 1/7 (14.3%) | 1/15 (6.7%) | 0/8 | 0/5 | 0/13 | 1/28 (3.6%) |
| Jaundice | 0/8 | 1/7 (14.3%) | 1/15 (6.7%) | 2/8 (25%) | 3/5 (60%) | 5/13 (38.5%) | 6/28 (21.4%) |
| Oliguria/decreased urination/anuria | 0/8 | 3/7 (42.9%) | 3/15 (20%) | 0/8 | 2/5 (40%) | 2/13 (15.4%) | 5/28 (17.9%) |
| Lymphadenopathy | 1/6 (16.7%) | 0/7 | 1/13 (7.7%) | 0/5 | 0/3 | 0/8 | 1/21 (4.8%) |
| Cardiac arrhythmia | 0/6 | 0/7 | 0/13 | 0/5 | 0/3 | 0/8 | 0/21 |

ED: emergency department, DPO: days post onset.

**TABLE 4:** Treatment and outcomes during the ED visit and hospitalization for admitted patients among leptospirosis cases identified through hospital-based active surveillance in Puerto Rico, 2019-2021.

| Leptospirosis cases |  |  |  |  |  |  |  |
| --- | --- | --- | --- | --- | --- | --- | --- |
|  | Probable |  |  | Confirmed |  |  | Leptospirosis Total |
|  | ED surveillance | Lab-Referral | Probable Total | ED surveillance | Lab-Referral | Confirmed Total |  |
| Treatment during ED and hospitalization |  |  |  |  |  |  |  |
| Abx prescribed at ED visit or hospitalization | 4/8 (50%) | 7/7 (100%) | 11/15 (73.3%) | 7/8 (87.5%) | 5/5 (100%) | 12/13 (92.3%) | 23/28 (82.1%) |
| More than one Abx prescribed | 3/4 (75%) | 5/7 (71.4%) | 8/11 (72.7%) | 2/7 (28.6%) | 2/5 (40%) | 4/12 (33.3%) | 12/23 (52.2%) |
| Intubation/ventilation | 0/8 | 0/7 | 0/15 | 0/8 | 1/5 (20%) | 1/13 (7.7%) | 1/28 (3.6%) |
| Supplemental oxygen | 1/8 (12.5%) | 2/7 (28.6%) | 3/15 (20%) | 1/8 (12.5%) | 1/5 (20%) | 2/13 (15.4%) | 5/28 (17.9%) |
| Blood/blood product transfusion | 0/8 | 0/7 | 0/15 | 0/8 | 2/5 (40%) | 2/13 (15.4%) | 2/28 (7.1%) |
| Vasopressors | 0/8 | 1/7 (14.3%) | 1/15 (6.7%) | 0/8 | 1/5 (20%) | 1/13 (7.7%) | 2/28 (7.1%) |
| Disposition after ED visit |  |  |  |  |  |  |  |
| Discharged from ED | 6/8 (75%) | 0/7 | 6/15 (40%) | 4/8 (50%) | 0/5 | 4/13 (30.8%) | 10/28 (35.7%) |
| Transferred other hospital | 0/8 | 0/7 | 0/15 | 0/8 | 2/5 (40%) | 2/13 (15.4%) | 2/28 (7.1%) |
| Admitted same hospital | 2/8 (25%) | 7/7 (100%) | 9/15 (60%) | 4/8 (50%) | 3/5 (60%) | 7/13 (53.9%) | 16/28 (57.1%) |
| Hospitalization days average (range) | 5 (4 – 6) | 4.1 (2-8) | 4.3 (2-8) | 5.5 (5-7) | 6 (1-9) | 5.7 (1-9) | 4.9 (1-9) |
| Final outcome |  |  |  |  |  |  |  |
| Full symptom recovery<sup>1,2</sup> | 5/8 (62.5%) | 3/7 (42.9%) | 8/15 (53.3%) | 4/6 (66.7%) | 1/4 (25%) | 5/10 (50%) | 13/25 (52%) |

ED: emergency department
<sup>1</sup> Ascertained based on a convalescent interview for 25 patients conducted on average 26 days after the ED visit.
<sup>2</sup> There were no deaths among leptospirosis cases based on information at discharge for three cases and convalescent interview for 25 cases.

### 3.4 Clinical laboratory examinations at the ED visit

All confirmed and probable cases had clinical laboratory tests completed but the types of tests ordered by the attending physician varied. The most common clinical laboratory blood tests ordered among all cases were complete blood count (CBC), blood urea nitrogen (BUN), creatinine, electrolytes, and glucose (27/28, 96.4% each) (Table S2). The most common laboratory findings among confirmed cases were thrombocytopenia (10/12, 83.3%), high AST (8/10, 80%), hyperglycemia (11/13, 84.6%), high bilirubin (6/8, 75%), high BUN (8/13, 61.5%), high ALT (6/10, 60%), and low albumin (5/9, 55.6%). Abnormal laboratory findings were less common among probable cases, with elevated AST (6/8, 75%) and hyperglycemia (9/14, 64.3%) being the most frequent findings. Urinalysis was ordered for 75% (21/28) of all cases, including for 92% (12/13) of confirmed cases, and for 60% (9/15) of probable cases. Proteinuria (13/21, 62%) and hematuria (13/21, 62%) were the most frequent findings among all cases with urinalysis done. These two findings were particularly frequent among confirmed cases with 83% (10/12) having proteinuria and 75% (9/12) having hematuria, compared with 33.3% (3/9) proteinuria and 44.4% (4/9) hematuria among probable cases (Table S2).

### 3.5 EMR diagnosis and laboratory infectious disease testing after ED or hospital discharge

Twenty-five of the 28 cases (89.3%) had a diagnosis reported in the EMR, with the number of entries ranging from one to four (Table S3). Leptospirosis was a diagnosis in 32.1% (9/28) of all cases, including 38.5% (5/13) of confirmed cases and 26.7% (4/15) of probable cases. All five (5/5, 100%) laboratory-positive referral confirmed cases and three of the seven (42.9%) laboratory-positive referral probable cases had a diagnosis entry of leptospirosis. Among the ED- enrolled cases, only one probable case (1/8, 12.5%) and none of the confirmed cases (0/8) received a leptospirosis diagnosis at the time of the ED visit or discharge from hospitalization. The most common non-leptospirosis diagnoses among all cases were viral syndrome (7/28, 25%), acute kidney injury (AKI) (4/28, 14.3%), and urinary tract infection (UTI) (4/28, 14.3%). Viral syndrome and UTI diagnoses were more common among ED-enrolled cases (7/16, 43.8% and 3/16, 18.8%, respectively) than among laboratory-positive referral cases (0/12, 0% and 1/12, 8.3%, respectively). AKI was more common among laboratory-positive referral cases (3/12, 25%) than ED-enrolled cases (1/16, 6.3%). Acute liver failure was diagnosed in one patient, a laboratory-positive referral confirmed case. Other diagnoses included dengue, mycoplasma, sepsis, upper respiratory infection, COVID-19, and unspecified fever (Table S4). The proportion of cases with non-leptospirosis infectious disease laboratory diagnostics conducted was high (25/28, 89.3%) and driven largely by Mycoplasma, Influenza, and COVID-19 test orders (Table S3). Testing resulted in one case (a confirmed case) positive for Influenza B (5.9%, 1/17), two cases (one probable and one confirmed) positive for Mycoplasma (12.5%, 2/16), and one case (a probable case) positive for COVID-19 (5.3%, 1/19). One confirmed case had a clinical diagnosis of dengue in the EMR, without supporting laboratory test results. The only dengue test ordered was for a patient with a diagnosis of unspecified fever with a negative test result. A hepatitis test was ordered for four case-patients with negative results.

### 3.6 Treatment prior to and during the ED visit and hospitalization

Patient history revealed that 42.9% (12/28) of the cases sought care prior to the enrollment visit, including a greater proportion of laboratory-positive referrals (58.3%, 7/12) than ED-enrolled cases (31.3%, 5/16) (p = 0.152). Four of them (33.3%, 4/12), all laboratory-positive referral cases, had been taking antibiotics for the current illness. Two ED-enrolled cases reported taking antibiotics but did not report seeking care (Table 3). Among all cases, 82.1% (23/28) were prescribed antibiotics at the time of the enrollment visit, including those subsequently hospitalized, and the most common antibiotics prescribed were ceftriaxone (73.9%, 17/23), piperacillin (30.4%, 7/23), and doxycycline (26.1%, 6/23) (Table S5). Antibiotic prescription was 92.3% (12/13) for confirmed patients and 73.3% (11/15) for probable cases (p = 0.333) (Table 4). The most common antibiotics prescribed among confirmed cases were ceftriaxone (75%, 9/12) and doxycycline (33.3%, 4/12). A wider range of antibiotics were prescribed for probable cases (Table S5). All laboratory-positive referral cases (12/12, 100%) were prescribed antibiotics compared with 68.8% (11/16) of ED-enrolled cases (p = 0.053) (Table 4). Supplemental oxygen (5/28, 17.9%) was the most common non-antibiotic treatment among all cases at any point during presentation and/or hospitalization. A few cases received a blood transfusion (2 confirmed cases), vasopressors (one probable and one confirmed case), and intubation/ventilation (one confirmed case) (Table 4). No leptospirosis cases received dialysis.

### 3.7 Disposition after the ED visit

After the ED visit, 64.3% (18/28) of the cases required hospitalization, 11.1% (2/18) were transferred to another hospital and 88.9% (16/18) were admitted to the same hospital. Hospitalization was required for 37.5% (6/16) of ED-enrolled cases and for 100% (12/12) of laboratory-positive referral cases (p < 0.001). Two (12.5%, 2/16) of the cases admitted to the same hospital were admitted to the ICU, both were confirmed leptospirosis cases. The overall average duration of hospitalization was 4.9 days; with an average of 4.3 days for probable cases and 5.7 days for confirmed cases (p = 0.220) and an average of 5.3 days for ED-enrolled cases and 4.7 days for laboratory-positive referral cases (p = 0.477) (Table 4). Among the 16 patients admitted to the same hospital, 15 (93.8%) were discharged, and one (a laboratory-positive referral probable case) was transferred to another hospital after 2 days.

A total of 25 cases (25/28, 89.3%), 10 confirmed and 15 probable, completed the convalescent questionnaire administered by project personnel on average 29.5 days (range: 11- 111 days) after illness onset. Overall, 52% (13/25) of the interviewed patients reported full recovery at the time of the interview, including 50% (5/10) and 53.3% (8/15) of confirmed and probable cases, respectively. Fewer laboratory-positive referral cases reported full recovery (4/11, 36.4%) compared with ED-enrolled cases (9/14, 64.3%) (Table 4). Among those cases that were interviewed ≤30 DPO, 41.2% (7/17) reported full recovery. Persisting clinical symptoms reported at the follow-up interview by cases not fully recovered included headache in 8.3% (2/12) of cases, pain (abdominal, calf, muscle, joint) in 66.7% (8/12), fatigue (tiredness, weakness) in 41.7% (5/12), gastrointestinal symptoms (diarrhea, nausea, vomiting) in 25% (3/12), shortness of breath in 25% (3/12), and light sensitivity in 8.3% (1/12). Overall, the proportion of cases reporting full recovery was 50% (10/20) for those who were prescribed antibiotics at the ED visit and 60% (3/5) for those who were not. Among those interviewed ≤30 DPO, these proportions were 42.9% (6/14) and 33.3% (1/3), respectively. Four of the 25 cases (16%) stated that a caretaker had to miss work or school to care for the patient during the illness and for an average of 4.8 days (range: 1-8 days). Among 13 cases who responded to the question, the average days they missed school or work due to the illness was 8 days (range: 0-24 days). Based on data collected at the time of discharge for 3 patients and the convalescent interview for the remaining 25 patients, there were no deaths among the leptospirosis cases identified.

### 3.8 Clinical presentation of leptospirosis cases in relation to the CSTE definition

Overall, all but one of the leptospirosis cases (27/28, 96.4%) met at least one of the CSTE definitions for clinical compatibility with leptospirosis. One probable case, enrolled through the ED, did not meet either of the two groups of clinical criteria; this patient’s chief complaints were lower back, joint,and neck pain, and the patient was diagnosed with lumbago. The CSTE AFI criterion was present in 71.4% (20/28) of all cases; 84.6% (11/13) of confirmed cases and 60% (9/15) of probable cases. The CSTE Severe criterion was met by 89.3% (25/28) of all cases, 92.3% (12/13) of confirmed cases, and 86.7% (13/15) of probable cases. When using patients classified as not a case and fever patients combined as the comparison group, the SE and SP for the CSTE AFI criteria to identify patients with leptospirosis was 71.4% (95% C.I.: 51.3%, 86.8%) and 40.2% (95% C.I.: 35.3%, 45.2%), respectively. The SE for the CSTE Severe criteria was 89.3% (95% C.I.: 71.8%, 97.7%) and the SP was 8.2% (95% C.I.: 5.7%, 11.4%). For meeting either CSTE criteria, SE was 96.4% (95% C.I.: 81.7%, 99.9%), and SP was 5.6% (95% C.I.: 3.6%, 8.4%). Figure 3 shows the proportion of specific symptoms for leptospirosis cases (confirmed and probable) compared to all other enrolled patients (those classified as not a case and fever patients combined). Grouping symptoms into the same diagnosis/syndromes used for the CSTE definition showed that GI symptoms (22/28, 78.6% vs. 268/391, 68.5%), hemorrhagic symptoms (8/28, 28.6% vs. 35/391, 9%) and AKI with jaundice (3/28, 10.7% vs. 2/391, 0.5%) were more prevalent among leptospirosis cases compared with all other enrolled patients. Conversely, pulmonary symptoms were more prevalent among fever patients and those classified as not a case (246/391, 62.9%) than among leptospirosis cases (13/28, 46.4%) (Table 5). The prevalence of leptospirosis cases was 3.5 times greater among patients with hemorrhagic symptoms compared to those without (PR = 3.5, 95% C.I.:1.6, 7.5). Patients with AKI and jaundice had a 9.9 times greater prevalence of leptospirosis cases compared to patients without (PR = 9.9, 95% C.I.: 4.4, 22.3) (Table 5).

**FIGURE 3:**
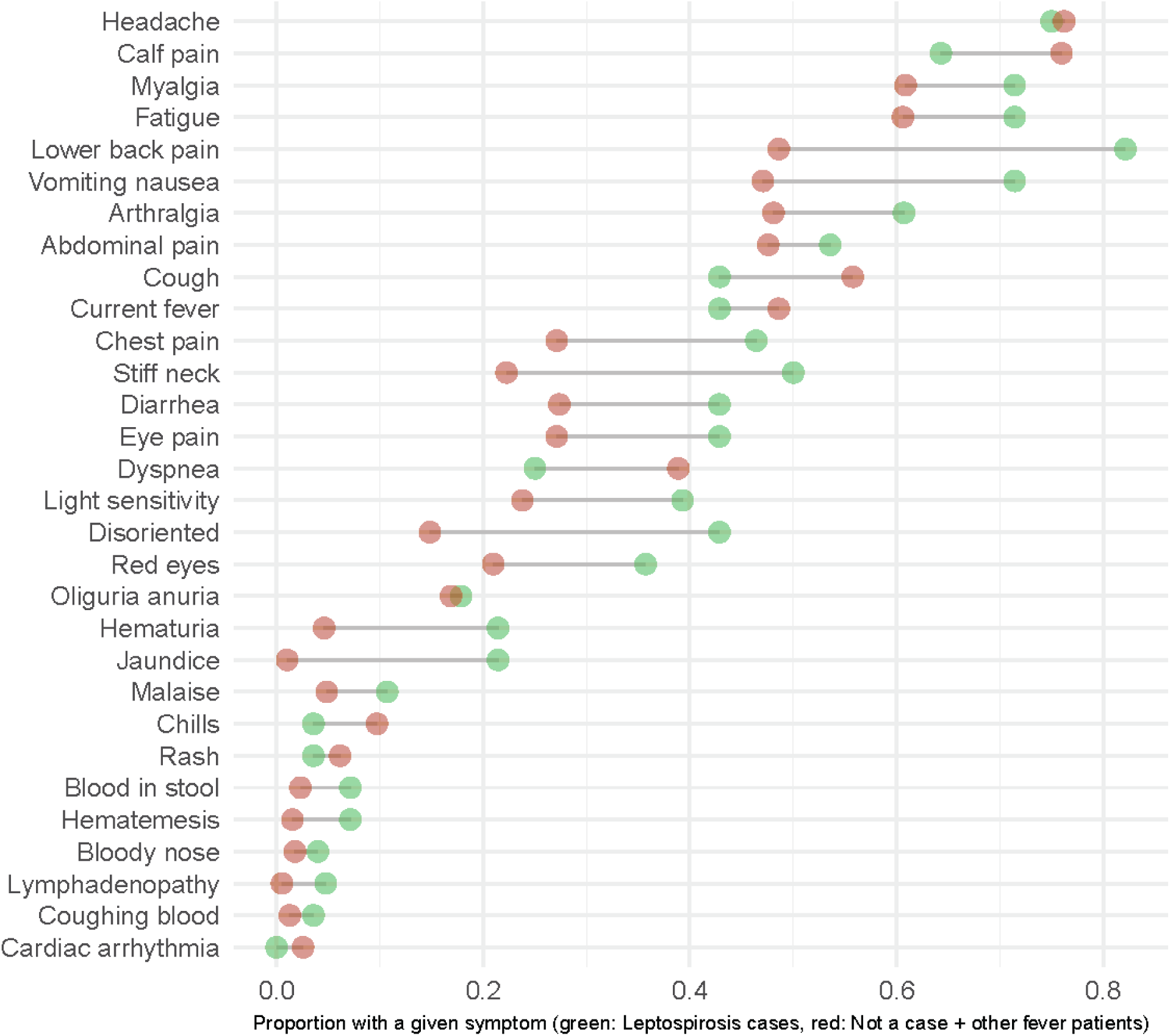
Proportion of a given symptom at the time of hospital emergency department visit among patients enrolled in active surveillance and classified as confirmed or probable leptospirosis cases (n = 28) and those classified as not a case or other fever patients combined (n = 391), Puerto Rico, 2019-2021.

**TABLE 5:**
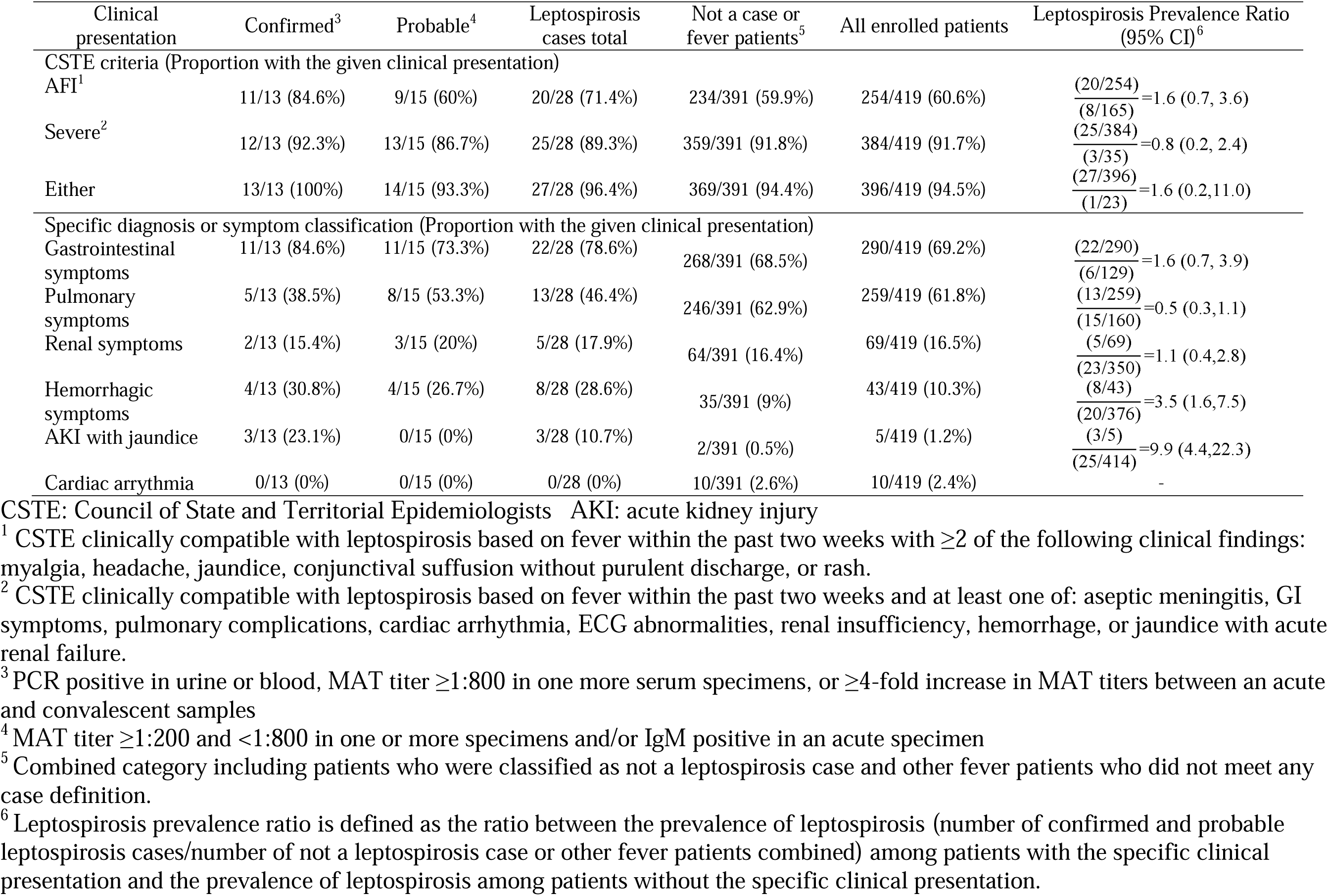
Evaluation of the CSTE clinical criteria for leptospirosis based on confirmed and probable leptospirosis cases identified in active hospital-based surveillance in Puerto Rico, 2019-2021.

## 4. Discussion

UAFI is one of the most common reasons for patients seeking healthcare worldwide and is caused by a wide range of etiologies [32]. Enrollment of patients among those seeking care at an ED who met the criteria of UAFI and testing everyone for leptospirosis using a standard diagnostic algorithm, regardless of clinical presentation and/or exposure history, allowed unbiased diagnostic efforts and identification of some cases that otherwise may not have been detected. In this investigation, we demonstrated a 4% prevalence of leptospirosis cases (probable and confirmed) among the ED-enrolled patients with UAFI. The proportion of leptospirosis-attributed etiology among febrile patients is expected to vary according to geographical and seasonal variations in differing eco-epidemiological conditions. Making meaningful prevalence comparisons across different places and times has the additional challenge of different studies using diverse inclusion and exclusion criteria, populations for case-finding, diagnostic testing protocols, and case definitions. For example, a study in Peru identified 9% leptospirosis confirmed cases among hospital patients presenting with fever of ≤5 days and other non-specific symptoms and using MAT as the only diagnostic test[33]. A study in New Zealand enrolled hospital patients based on the presentation of non-specific febrile illness symptoms for ≤4 days and identified 16% with confirmed or probable leptospirosis using MAT [34]. Additional studies investigated leptospirosis etiology among patients with other presumptive diagnoses such as a study in Colombia using paired serum samples from presumptive dengue patients that identified 24% as confirmed leptospirosis cases using MAT [35]. In Thailand, a 4% prevalence of confirmed leptospirosis cases was identified using an IgM rapid test, MAT, and PCR, among UAFI patients testing negative for other infectious etiologies and excluding those with obvious single organ-specific symptoms or signs of a non-leptospirosis infection [36]. A study in Sri Lanka estimated prevalences of leptospirosis confirmed cases from 16% to 45%, depending on the hospital and year, among UAFI patients after excluding lower respiratory infections and meningitis [37]. These different approaches for active surveillance can have a significant impact on the estimate of the percentage of febrile illness attributed to leptospirosis. Using more restrictive criteria, such as enrolling patients in only the first few days of illness and/or with current fever (but not recent history of fever) will miss leptospirosis cases that delay seeking care. Including patients based on specific symptoms or exposure history that align with more typical leptospirosis presentations, or excluding them because of organ-specific infections can reduce the number of potential patients that need to be tested but can lead to missed cases and underestimation of prevalence, as well as inaccurate characterization of risk factors. Additionally, the choice of laboratory testing will greatly impact the ability to detect cases as using multiple tests, especially serologic and molecular tests in parallel, can increase the probability of case-detection. In our investigation, we used purposefully broad UAFI criteria to obtain a less biased estimate of leptospirosis prevalence, allowing us to characterize the wide spectrum of clinical presentations associated with the infection. Importantly, it is essential to conduct active surveillance for leptospirosis in the geographic area of interest, as it is known that leptospirosis incidence and local clinical and epidemiological characterization can vary significantly between different locations and climates. We conducted our investigation in the years following the 2017 Hurricanes Irma and Maria since our goal was to better characterize the clinical presentation and epidemiology of leptospirosis in Puerto Rico in order to better prepare for future disasters.

The use of PCR was crucial for identifying and promptly confirming leptospirosis cases among ED-enrolled patients. Although not readily available in all healthcare settings, there is consensus on the benefits of PCR as a tool to provide a rapid and reliable diagnosis, especially during the early stages of the infection when patients may be negative by serological tests [37, 38]. However, combining PCR with serologic testing is very important since leptospires only remain in the blood for a few days after infection. The use of commercially available IgM rapid tests is increasing and serves as a more accessible tool than PCR because they do not need highly specialized equipment; however, diagnostic efforts relying on IgM tests alone would miss cases due to low sensitivity during the first few days to a week of illness. As revealed by our results, of the eight PCR positive cases found through ED-enrollment, five were IDOT IgM negative at the acute visit and would have been missed based on IgM testing alone. Conversely, our additional enrollment of clinically suspected cases with a routinely ordered IDOT IgM borderline or positive result (laboratory-positive referrals) allowed further laboratory testing of 13 patients. MAT serology allowed confirmation of five of these cases, which were enrolled on average 10 DPO (Table 2), after which MAT is expected to have high sensitivity [39]. Overall, findings in this surveillance investigation underscore the need for a combined diagnostic effort using local PCR and IgM testing to maximize the likelihood of early and prompt case detection with follow-up MAT antibody testing to support additional serologic confirmation and epidemiological surveillance.

The majority of cases identified were men and of low income which are commonly reported characteristics of leptospirosis cases. Case-history showed that more than half of the reported occupations were classified as homemaker, student, retired, unemployed, or a service/retail type of employment, which are not traditionally considered high-risk groups for leptospirosis. Historically, commonly reported at-risk occupations are those such as animal caretakers, sewer workers, and those with fresh water and other outdoor exposures [3]. Contact with mud or wet soil, the occurrence of recent heavy rain, and the presence of rodents and dogs were common among the cases identified (Table S1), and are specific factors that could expose people to *Leptospira* bacteria through routine daily activities. The case demographics and potential exposures in this investigation are consistent with previous descriptions of leptospirosis cases in Puerto Rico, such as the high frequency of cases who were males, retirees, or students [14] and factors that could lead to environmental exposure, such as through heavy rain [14] and living in proximity to surface freshwater sources (i.e., canal) [40]. These case profiles highlight the importance of obtaining a complete intake history on patients presenting with UAFI to prompt the inclusion of leptospirosis as a diagnostic differential and initiate any follow-up surveillance investigations needed. Additionally, the three *Leptospira* species isolated from the cases have been identified in animal reservoirs in Puerto Rico; *L. interrogans* in rodents, dogs, and mongoose, *L. borgpetersenii* in rodents and cattle, and *L. kirschneri* in rodents [41, 42]. Future studies are needed to elucidate specific transmission pathways and inform control programs.

A task force synthesis report on leptospirosis clinical presentation describes fever, rigors, myalgia, headache, and conjunctival suffusion as commonly present at the early or leptospiremic phase of the illness while cough, nausea, vomiting, and diarrhea are present with less frequency [43]. Some patients develop a more severe illness and can present arrhythmias, ocular pain, lymphadenopathy, hepatosplenomegaly, skin rash, aseptic meningitis and less often arthralgias, bone pain, sore throat, and abdominal pain [43]. Leptospirosis patients who develop the more recognized Weill’s disease show a presentation consistent with renal and hepatic failure, and in some cases, pulmonary hemorrhage [44]. Consistent with previous reports, among the cases identified in this surveillance investigation, the most common symptoms (over 50% of all cases) included myalgia, headache, vomiting/nausea, lower back pain, fatigue/weakness, calf pain, abdominal pain, stiff neck, and arthralgia. Red eyes, cough, and diarrhea were also reported but at a slightly lower frequency. There were no reports of arrhythmia or meningitis among the cases; however, these are part of recognized possible presentations with varying frequencies depending on the study [45–47] and detection in association with leptospirosis may need a high level of clinical suspicion if classical signs are not present[48]. Fever is a fundamental component of UAFI and was a primary inclusion criterion for this surveillance investigation. All enrolled cases had either a current fever or a history of fever in the past two weeks. Notably, only 43% of all cases exhibited measured fever at the time of the ED visit. Other investigations commonly report much higher occurrences of fever [49–51]. However, comparisons are limited due to the utilization of different time periods in the inclusion criteria (e.g., fever in the past 3 days or 2 weeks prior to the visit), a different set of clinical symptoms for the inclusion criteria, as well as a tendency to not report current fever and history of fever separately. Nevertheless, this underscores the importance of obtaining a fever history at intake and the knowledge that a fever at the time of presentation is not required to be a suspected case of leptospirosis. The largely nonspecific presentations for many cases typically make leptospirosis indistinguishable from other UAFI etiologies based on clinical presentation alone.

The classic presentation of leptospirosis illness that includes renal and hepatic damage with or without unexplained hemorrhage presents with abnormal clinical laboratory results and is often indicative of illness severity. A previous investigation in Puerto Rico of a concurrent dengue and leptospirosis outbreak in 2010 reported 15% fatality among 175 identified leptospirosis cases and found that death among leptospirosis cases was associated with decreased serum bicarbonate combined with thrombocytopenia, and increased WBC count combined with elevated creatinine [16]. The frequencies of high WBC and high creatinine were generally low among the cases in this surveillance investigation (Table S2) and only two cases presented with both (7.7%); however, thrombocytopenia, which could be a contributing factor to hemorrhages [43, 52] and hyperbilirubinemia, the result of hepatocellular damage and disruption of hepatocyte intercellular junctions were common among confirmed cases and particularly among laboratory-positive referral confirmed cases (Table S2). The presence in this enrollment group of these altered clinical markers, along with clinical signs indicative of more complicated illness is consistent with clinical suspicion of leptospirosis by the clinician who ordered the routine leptospirosis IDOT IgM diagnostic test. In this surveillance investigation, there were no deaths among the cases up to the point of the last encounter (discharge or convalescent survey) but a high percentage (64%) required hospitalization and 13% required ICU admission. However, this investigation was likely biased towards enrolling leptospirosis cases with more severe presentations as it was conducted at hospitals, and therefore patients with milder illness for which they did not seek medical care or went to a local clinic would not have been captured. Consistent with their more complicated presentations, these hospitalization rates were driven by confirmed and laboratory positive referral cases. Comparing hospitalization and ICU admission rates between studies is difficult due to variable case definitions and enrollment methods, including some in which identification of patients is based on leptospirosis clinical suspicion and would likely enroll patients that may present classic, and potentially more severe, leptospirosis presentation, consequently requiring hospitalization. A sentinel hospital surveillance investigation in Sri Lanka enrolling hospitalized patients had a 7% ICU admission and 3% fatality [49], and a report of mostly hospitalized patients from the Colombia leptospirosis surveillance program estimated a 30% ICU admission and 9% fatality [45]. These and other leptospirosis case investigations are primarily based on clinically suspected and hospitalized cases, which limits the observation of clinical presentation across the illness spectrum, especially in milder cases, and also limits the evaluation of risk factors for hospitalization or severe outcomes. We have insufficient data to perform an evaluation of patient factors associated with clinical severity; however, few or no cases presented with complications leading to the most severe and fatal outcomes such as shock, meningitis, cardiac or pulmonary involvement, or need for dialysis [46] which may have contributed to no fatalities being observed.

This surveillance investigation tracked project-requested leptospirosis diagnostics separately from routine care and EMR diagnosis entries, although project diagnostic results were provided to the hospitals; therefore, the low number of EMR leptospirosis diagnoses recorded among identified confirmed and probable leptospirosis cases reflects the challenges of EMR data utilization for clinical and epidemiological leptospirosis research and surveillance due to underdiagnosis. Laboratory diagnostic test utilization for other diseases was high (89%), mostly for common respiratory etiologies such as influenza, Mycoplasma, and COVID-19, and resulted in positive test results in three identified leptospirosis cases (Table S4) that may indicate co-infections. Concurrent febrile illness etiologies are expected at the population level, however, there is limited research on the prediction, detection, and impact of acute co-infections. Viral syndrome was a common EMR diagnosis among the cases detected, particularly for ED-enrolled cases, which happens when viral infections are predominant in the populations and are therefore commonly attributed as the likely cause of the febrile syndromes. In this case, the consequences of an incorrect viral syndrome diagnosis would be detrimental to providing appropriate and timely antibiotic treatment for patients with leptospirosis. Some mild leptospirosis infections can be self-limiting, but clinical guidelines recommend initiating antibiotic treatment promptly after clinical suspicion of leptospirosis to improve prognosis, shorter illness duration, and improved symptoms [53]. Doxycycline is a drug of choice for mild symptoms, and alternative options include ampicillin, amoxicillin, or azithromycin. Intravenous penicillin is the drug of choice for patients with severe leptospirosis; ceftriaxone and cefotaxime are alternative antimicrobial agents [54]. In cases identified, the practice of starting antibiotic treatment immediately upon suspicion of leptospirosis was consistently applied to the laboratory-positive referral cases, and the one ED-enrolled case with an EMR clinical leptospirosis diagnosis recorded as 100% received antibiotics, with ceftriaxone as the most common drug. The majority of other ED-enrolled cases were prescribed antibiotics (Table S5), despite having EMR diagnoses of viral syndrome, unspecified fever, or lack of a confirmed bacterial etiology. While this may have benefited the illness evolution, it highlights the challenges in febrile illness clinical differentiation and management.

A leptospirosis case definition is important for identifying cases and standardizing reporting and analysis of surveillance data. The clinical presentation component of the CSTE definition is developed based on the most current understanding of the disease to identify a set of clinical criteria compatible with leptospirosis. Most cases, including all confirmed cases, satisfied the CTSE clinical criteria (Table 5). This is expected as the criteria encompass a comprehensive range of non-specific signs and symptoms associated with leptospirosis, from mild to severe manifestations. Because of this same reason, it was also expected that a considerable proportion of the remainder of the febrile illness patients (not identified as leptospirosis cases) met the CSTE clinical criteria, highlighting the need to pursue laboratory testing to determine a diagnosis. This finding is reflected in the high SE calculated for the CSTE clinical criteria and the very low SP. Warnasekara et al. evaluated a slightly narrower surveillance case definition among undifferentiated febrile illness patients in Sri Lanka and found a similarly low SP [37]. This lack of SP for the clinical criteria means that in Puerto Rico and similar settings where many other possible diseases that mimic leptospirosis are circulating, many febrile illness patients would meet the clinical criteria compatible with leptospirosis but have a different etiology. Obtaining an accurate burden estimation for leptospirosis requires, among other approaches, a large emphasis on laboratory testing, along with the corresponding allocation of resources. There have been efforts to improve the clinical criteria used to identify presumptive leptospirosis cases using scoring algorithms based on epidemiological features, specific clinical signs and/or clinical laboratory results [55–59], however, there has been limited success in establishing clinicaly useful guidelines for leptospirosis differentiation from other causes of UAFI without supplemental diagnostic testing results. In our Puerto Rico setting, presenting with hemorrhagic symptoms or with AKI together with jaundice were significantly associated with being classified as a leptospirosis case (Table 5) and would be important clinical variables to consider in the development of locally relevant case-identification guidelines. In general, further research is needed to identify a set of optimal criteria for identifying presumptive leptospirosis patients, which may vary by population and the locally circulating *Leptospira* strains, and to ensure equitable access to laboratory resources.

Several limitations in this surveillance investigation include the small sample size that limited a more comprehensive analysis of factors associated with hospitalization or severity. Non-leptospirosis laboratory testing, including basic metabolic profiles, were ordered for a fraction of the cases, at the discretion of the clinician, which limits the interpretation across different categories of enrolled patients. We applied a standardized diagnostic algorithm using commonly recommended tests and interpretations; however, laboratory tests are imperfect and there could still be missed leptospirosis cases among the febrile illness patients tested due to low concentration of analyte and/or early antibiotic administration that could thwart the immune response. Alternatively, IDOT IgM could result in false positive results depending on the prevalence of cross-reacting infections, for example, *Salmonella typhi*. Leptospirosis cases identified were among those seeking care at the ED and at a small number of hospitals in Puerto Rico, and, therefore, would not include case characteristics for those seeking care at primary care clinics or not seeking medical care at all, which likely represent cases with milder clinical presentations.

## 5. Conclusion

This project reports the fraction of leptospirosis probable or confirmed cases among acute febrile illness patients seeking care in the emergency room of selected hospitals in Puerto Rico. Because of the broad inclusion criteria, the estimated 4% of cases is an important finding considering the diverse origins of fever etiologies and the commonly large burden of respiratory infections. The broad spectrum of leptospirosis clinical presentations was demonstrated and the significant role of comprehensive diagnostic testing in identifying leptospirosis cases was emphasized. The need to increase awareness among clinicians about including leptospirosis in the differential diagnosis of UAFI was highlighted. The data contribute insights into the characteristics of cases identified independently of clinician suspicion compared with those identified through routine practices. While all the cases received antibiotic treatment and were discharged after the hospital ED visit or hospitalization, results revealed that further research is needed to optimize early diagnosis, inform treatment strategies, and ensure favorable and full recovery after the acute leptospirosis illness.

## Data Availability

All data produced in the present study are available upon reasonable request to the authors.

## Data Availability

The data that support the findings of this study are available on request from the corresponding author. The data are not publicly available due to privacy or ethical restrictions.

## Authors’ Contributions

Conceived the overall project and funding (CMZ, IS, AK), project management and implementation (CMZ, AK, MM, IS), project execution and operations (CMZ, AK, MM) contribution to methodology and various elements of data collection (CMZ, MM, IS, NCD, RRG, LSR, CRC, RMR, RC, NPC, DRN, JMA, RG, DH, LM, RRO, HW), data curation (CMZ, MM, CRC, NCD), concept of this paper, data analysis, and wrote original draft of the manuscript (CMZ). All authors reviewed and approved the final version of the manuscript.

The findings and conclusions in this report are those of the authors and do not necessarily represent the official position of the Centers for Disease Control and Prevention.

## Acknowledgments

The authors would like to express their gratitude to the participating hospitals, including the administration, clinicians, nurses, IT personnel, and clinical laboratory personnel for their interest in the project and their contributions to the implementation of project procedures. We also extend our thanks to the patients for their willingness to enroll and provide specimens and data, and to Robyn Stoddard, DVM, PhD from the Centers for Disease Control and Prevention who performed part of the laboratory testing. Additionally, the authors want to acknowledge and thank the other hospitals that were visited during the site selection process for their interest in participation.

