## Supplemental Tables and Figures for "Diagnosis and clinical description of leptospirosis cases identified through hospital-based active surveillance in Puerto Rico, 2019-2021"

**Supplementary material**

TABLE S1: Patient demographic and risk factor characteristics among laboratory-confirmed and probable leptospirosis cases identified through hospital-based active surveillance, Puerto Rico, 2019-2021. Risk factors were identified in the 30 days before illness onset.

| Characteristics | Probable (n = 15) | Confirmed (n =13) | Total (n = 28) |
| --- | --- | --- | --- |
| Age (yrs): average (range) | 39.2 (6-81) | 47.7 (20-68) | 43.1 (6-81) |
| Sex: |  |  |  |
| Male | 8/15 (53.3%) | 12/13 (92.3%) | 20/28 (71.4%) |
| Race:^1^ |  |  |  |
| Black | 4/9 (44.4%) | 4/6 (66.7%) | 8/15 (53.3%) |
| White | 5/9 (55.6%) | 3/6 (50%) | 8/15 (53.3%) |
| American Indian /Alaskan native | 2/9 (22.2%) | 0/6 | 2/15 (13.3%) |
| Occupation: |  |  |  |
| Service/retail^2^ | 3/15 (20%) | 3/13 (23.1%) | 6/28 (21.4%) |
| Janitor | 1/15 (6.7%) | 0/13 | 1/28 (3.6%) |
| Gardener | 0/15 | 1/13 (7.7%) | 1/28 (3.6%) |
| Healthcare worker | 2/15 (13.3%) | 0/13 | 2/28 (7.1%) |
| Animal caretaker/farmer | 0/15 | 5/13 (38.5%) | 5/28 (17.9%) |
| Manual laborer | 2/15 (13.3%) | 1/13 (7.7%) | 3/28 (10.7%) |
| Homemaker/student/other^2^ | 9/15 (60%) | 5/13 (38.5%) | 14/28 (50%) |
| Education level: |  |  |  |
| High school | 10/15 (66.7%) | 10/13 (76.9%) | 20/28 (71.4%) |
| Technical school/college | 5/15 (33.3%) | 3/13 (23.1%) | 8/28 (8.6%) |
| Income: |  |  |  |
| ≤ USD $20,000^3^ | 3/5 (60%) | 4/6 (66.7%) | 7/11 (63.6%) |
| Drinking water source: |  |  |  |
| Piped water | 9/15 (60%) | 7/13 (53.9%) | 16/28 (57.1%) |
| Bottled water | 13/15 (20%) | 11/13 (84.6%) | 24/28 (85.7%) |
| Open well | 0/15 | 1/13 (7.7%) | 1/28 (3.6%) |
| Bathing water source: |  |  |  |
| Piped water | 15/15 (100%) | 12/13 (92.3%) | 27/28 (96.4%) |
| Open well | 0/15 | 1/13 (7.7%) | 1/28 (3.6%) |
| Other water contact: |  |  |  |
| River/moving water | 0/15 | 2/13 (15.4%) | 2/28 (7.1%) |
| Mud or wet soil | 5/15 (33.3%) | 7/13 (53.8%) | 12/28 (42.9%) |
| Other sources^4^ | 3/15 (20%) | 1/13 (7.7%) | 4/28 (14.3%) |
| Heavy rain: | 8/14 (57.1%) | 7/13 (53.8%) | 15/27 (53.5%) |
| Recent flooding: | 1/15 (6.7%) | 4/13 (30.8%) | 5/28 (17.9%) |
| Open wounds/cuts: | 1/15 (6.7%) | 6/13 (46.2%) | 7/28 (25%) |
| Rodent presence at home or workplace: | 7/13 (53.8%) | 12/13 (92.3%) | 19/26 (73.1%) |
| Dog contact: | 9/15 (60%) | 9/13 (69.2%) | 18/28 (64.3%) |
| Horse contact: | 2/15 (13.3%) | 2/13 (15.4%) | 4/28 (14.3%) |
| Livestock contact: | 1/15 (6.7%) | 4/13 (30.8%) | 5/28 (17.9%) |

^1^ Thirteen people declined to answer.

^2^ Service/retail included customer service staff and food servers. Other occupations included retired and unemployed.

^3^ Income was unknown for 17 cases. The highest reported income category was $20,001-$40,000

^4^ Other water included drinking water for pets, sea water, water for laundry, and hose water.

TABLE S2: Clinical laboratory diagnostic findings among confirmed and probable leptospirosis cases identified through active hospital-based surveillance in Puerto Rico, 2019-2021

|  | Probable leptospirosis cases | | | Confirmed leptospirosis cases | | |  |
| --- | --- | --- | --- | --- | --- | --- | --- |
|  | ED surveillance  (n = 8) | Lab-Referral  (n = 7) | Probable Cases Total  (n = 15) | ED surveillance  (n = 8) | Lab-Referral  (n = 5) | Confirmed Cases Total  (n = 13) | Total leptospirosis cases (n = 28) |
| **Blood analysis** |  |  |  |  |  |  |  |
| Leukocytosis | 1/8 (12.5%) | 1/7 (14.3%) | 2/15 (13.3%) | 2/8 (25%) | 1/4 (25%) | 3/12 (25%) | 5/27 (18.5%) |
| Thrombocytopenia | 1/8 (12.5%) | 2/7 (28.6%) | 3/15 (20%) | 6/8 (75%) | 4/4 (100%) | 10/12 (83.3%) | 13/27 (48.2%) |
| Low hematocrit | 1/8 (12.5%) | 2/7 (28.6%) | 3/15 (20%) | 1/8 (12.5%) | 1/4 (25%) | 2/12 (16.7%) | 5/27 (18.5%) |
| Low hemoglobin | 1/8 (12.5%) | 0/7 | 1/15 (6.7%) | 1/8 (12.5%) | 0/4 | 1/12 (8.3%) | 2/27 (7.4%) |
| Neutrophils left shift | 0/8 | 1/7 (14.3%) | 1/15 (6.7%) | 0/8 | 0/4 | 0/12 | 1/27 (3.7%) |
| High BUN | 0/7 | 2/7 (28.6%) | 2/14 (14.3%) | 4/8 (50%) | 4/5 (80%) | 8/13 (61.5%) | 10/27 (37%) |
| High Creatinine | 1/7 (14.3%) | 2/7 (28.6%) | 3/14 (21.4%) | 2/8 (25%) | 2/5 (40%) | 4/13 (30.8%) | 7/27 (25.9%) |
| High Bilirubin | 0/2 | 1/5 (20%) | 1/7 (14.3%) | 2/3 (66.7%) | 4/5 (80%) | 6/8 (75%) | 7/15 (46.7%) |
| High AST | 2/3 (66.7%) | 4/5 (80%) | 6/8 (75%) | 3/5 (60%) | 5/5 (100%) | 8/10 (80%) | 14/18 (77.8%) |
| High ALT | 0/3 | 3/5 (60%) | 3/8 (37.5%) | 2/5 (40%) | 4/5 (80%) | 6/10 (60%) | 9/18 (50%) |
| High ALP | 0/3 | 1/5 (20%) | 1/8 (12.5%) | 1/3 (33.3%) | 1/3 (33.3%) | 2/6 (33.3%) | 3/14 (21.4%) |
| Low albumin | 0/3 | 1/5 (20%) | 1/8 (12.5%) | 2/4 (50%) | 3/5 (60%) | 5/9 (55.6%) | 6/17 (35.3%) |
| Low total protein | 0/3 | 1/5 (20%) | 1/8 (12.5%) | 1/5 (20%) | 1 /4 (25%) | 2/9 (22.2%) | 3/17 (17.7%) |
| Normal or low CPK | 1/1 (100%) | 1/1 (100%) | 2/2 (100%) | 2/2 (100%) | - | 2/2 (100%) | 4/4 (100%) |
| Hypokalemia | 2/7 (28.6%) | 1/7 (14.3%) | 3/14 (21.4%) | 1/8 (12.5%) | 3/5 (60%) | 4/13 (30.8%) | 7/27 (25.9%) |
| Hyponatremia | 1/7 (14.3%) | 1/7 (14.3%) | 2/14 (14.3%) | 4/8 (50%) | 1/5 (20%) | 5/13 (38.5%) | 7/27 (25.9%) |
| Hypernatremia | 0/7 | 1/7 (14.3%) | 1/14 (7.1%) | 0/8 | 0/5 | 0/13 | 1/27 (3.7%) |
| Hyperglycemia | 5/7 (71.4%) | 4/7 (57.1%) | 9/14 (64.3%) | 8/8 (100%) | 3/5 (60%) | 11/13 (84.6%) | 20/27 (74.1%) |
| High CRP | 0/2 | 1/2 (50%) | 1/4 (25%) | 1/2 (50%) | 0/1 | 1/3 (33.3%) | 2/7 (28.6%) |
| High ESR | 0/2 | 1/2 (50%) | 1/4 (25%) | 1/1 (100%) | - | 1/1 (100%) | 2/5 (40%) |
| High Lactic acid | 0/2 | 2/4 (50%) | 2/6 (33.3%) | 1/3 (33.3%) | 1/1 (100%) | 2/4 (50%) | 4/10 (40%) |
| High PT | 1/2 (50%) | 1/2 (50%) | 2/4 (50%) | 0/1 | 0/2 | 0/3 | 2/7 (28.6%) |
| High PTT | 0/2 | 0/2 | 0/4 | 0/1 | 1/2 (50%) | 1/3 (33.3%) | 1/7 (14.3%) |
| High D-dimer | 1/1 (100%) | - | 1/1 (100%) | 1/1 (100%) | 1/1 (100%) | 2/2 (100%) | 3/3 (100%) |
| **Urinalysis** |  |  |  |  |  |  |  |
| Proteinuria | 2/4 (50%) | 1/5 (20%) | 3/9 (33.3%) | 7/8 (87.5%) | 3/4 (75%) | 10/12 (83.3%) | 13/21 (61.9%) |
| Pyuria | 1/4 (25%) | 1/5 (20%) | 2/9 (22.2%) | 2/8 (25%) | 3/4 (75%) | 5/12 (41.7%) | 7/21 (33.3%) |
| Hematuria | 2/4 (50%) | 2/5 (40%) | 4/9 (44.4%) | 5/8 (62.5%) | 4/4 (100%) | 9/12 (75%) | 13/21 (61.9%) |
| Low Specific gravity | 1/4 (25%) | 3/5 (60%) | 4/9 (44.4%) | 1/6 (16.7%) | 0/3 | 1/9 (11.1%) | 5/18 (27.8%) |
| High Specific gravity | 0/4 | 1/5 (20%) | 1/9 (11.1%) | 3/6 (50%) | 0/3 | 3/9 (33.3%) | 4/18 (22.2%) |

TABLE S3: Diagnosis specified in the electronic medical records at the time of the ED visit among confirmed and probable leptospirosis cases identified through active hospital-based surveillance in Puerto Rico, 2019-2021

|  | Probable cases |  |  | Confirmed cases |  |  |  |
| --- | --- | --- | --- | --- | --- | --- | --- |
|  | ED surveillance  (n = 8) | Lab-Referral  (n = 7) | Probable Cases Total  (n = 15) | ED surveillance  (n = 8) | Lab-Referral  (n = 5) | Confirmed Cases Total  (n = 13) | Total leptospirosis cases (n = 28) |
| Dengue | 0/8 | 0/7 | 0/15 | 1/8 (12.5%) | 0/5 | 1/13 (7.7%) | 1/28 (3.6%) |
| Leptospirosis | 1/8 (12.5%) | 3/7 (42.9%) | 4/15 (26.7%) | 0/8 | 5/5 (100%) | 5/13 (38.5%) | 9/28 (32.1%) |
| Mycoplasma | 0/8 | 1/7 (14.3%) | 1/15 (6.7%) | 1/8 (12.5%) | 0/5 | 1/13 (7.7%) | 2/28 (7.1%) |
| Sepsis | 0/8 | 1/7 (14.3%) | 1/15 (6.7%) | 0/8 | 1/5 (20%) | 1/13 (7.7%) | 2/28 (7.1%) |
| Viral syndrome | 3/8 (37.5%) | 0/7 | 3/15 (20%) | 4/8 (50%) | 0/5 | 4/13 (30.8%) | 7/28 (25%) |
| Acute kidney injury | 0/8 | 1/7 (14.3%) | 1/15 (6.7%) | 1/8 (12.5%) | 2/5 (40%) | 3/13 (23.1%) | 4/28 (14.3%) |
| Acute liver failure | 0/8 | 0/7 | 0/15 | 0/8 | 1/5 (20%) | 1/13 (7.7%) | 1/28 (3.6%) |
| Urinary Tract Infection | 2/8 (25%) | 1/7 (14.3%) | 3/15 (20%) | 1/8 (12.5%) | 0/5 | 1/13 (7.7%) | 4/28 (14.3%) |
| Upper respiratory infection | 1/8 (12.5%) | 0/7 | 1/15 (6.7%) | 0/8 | 0/5 | 0/13 | 1/28 (3.6%) |
| Covid-19 | 0/8 | 1/7 (14.3%) | 1/15 (6.7%) | 0/8 | 0/5 | 0/13 | 1/28 (3.6%) |
| Unspecified fever | 0/8 | 0/7 | 0/15 | 2/8 (25%) | 0/5 | 2/13 (15.4%) | 2/28 (7.1%) |

TABLE S4: Non-leptospirosis diagnostic infectious disease laboratory tests ordered at the ED visit and results among confirmed and probable leptospirosis cases identified through active hostpital-based surveillance in Puerto Rico, 2019-2021

|  | Probable |  |  | Confirmed |  |  |  |
| --- | --- | --- | --- | --- | --- | --- | --- |
|  | ED surveillance  (n = 8) | Lab-Referral  (n = 7) | Probable Cases Total  (n = 15) | ED surveillance  (n = 8) | Lab-Referral  (n = 5) | Confirmed Cases Total  (n = 13) | Total leptospirosis cases (n = 28) |
| Any ID diagnostic test ordered | 7/8 (87.5%) | 7/7 (100%) | 14/15 (93.3%) | 7/8 (87.5%) | 4/5 (80%) | 11/13 (84.6%) | 25/28 (89.3%) |
| Influenza A | 0/5 | 0/3 | 0/8 | 0/5 | 0/4 | 0/9 | 0/17 |
| Influenza B | 0/5 | 0/3 | 0/8 | 1/5 | 0/4 | 1/9 (11.1%) | 1/17 (5.9%) |
| Strep | - | - | - | - | - | - | - |
| Mycoplasma | 0/3 | 1/4 (25%) | 1/7 (5.9%) | 1/5 (20%) | 0/4 | 1/9 (11.1%) | 2/16 (12.5%) |
| RSV | - | - | - | - | - | - | - |
| Dengue | - | - | - | 0/1 | - | 0/1 | 0/1 |
| Chikungunya | - | - | - | - | - | - | - |
| Zika | - | - | - | - | - | - | - |
| Hepatitis | 0/1 | 0/1 | 0/2 | 0/1 | 0/1 | 0/2 | 0/4 |
| COVID-19 | 0/4 | 1/6 (16.7%) | 1/10 (10%) | 0/5 | 0/4 | 0/9 | 1/19 (5.3%) |

TABLE S5: Antibiotics prescribed during the emergency room visit or hospitalization to leptospirosis cases identified through hospital-based active surveillance, Puerto Rico, 2019-2021

|  | Probable cases | | | Confirmed cases | | |  |
| --- | --- | --- | --- | --- | --- | --- | --- |
|  | ED surveillance | Lab-Referral | Probable Cases Total | ED surveillance | Lab-Referral | Confirmed Cases Total | Total leptospirosis cases |
| Doxycycline | 1/4 (25%) | 1/7 (14.3%) | 2/11 (18.2%) | 3/7 (42.9%) | 1/5 (20%) | 4/12 (33.3%) | 6/23 (26.1%) |
| Amoxicillin | 0/4 | 1/7 (14.3%) | 1/11 (9.1%) | 0/7 | 0/5 | 0/12 | 1/23 (4.4%) |
| Azithromycin | 1/4 (25%) | 2/7 (28.6%) | 3/11 (27.3%) | 0/7 | 0/5 | 0/12 | 3/23 (13%) |
| Ceftriaxone | 2/4 (50%) | 6/7 (85.7%) | 8/11 (72.7%) | 4/7 (57.1%) | 5/5 (100%) | 9/12 (75%) | 17/23 (73.9%) |
| Vancomycin | 1/4 (25%) | 1/7 (14.3%) | 2/11 (18.2%) | 0/7 | 0/5 | 0/12 | 2/23 (8.7%) |
| Piperacillin | 2/4 (50%) | 3/7 (42.9%) | 5/11 (45.5%) | 1/7 (14.3%) | 1/5 (20%) | 2/12 (16.7%) | 7/23 (30.4%) |
| Levofloxacin | 0/4 | 0/7 | 0/11 | 1/7 (14.3%) | 1/5 (20%) | 2/12 (16.7%) | 2/23 (8.7%) |
| Ciprofloxacin | 0/4 | 0/7 | 0/11 | 1/7 (14.3%) | 0/5 | 1/12 (8.3%) | 1/23 (4.4%) |
| Other cephalosporin^2^ | 1/4 (25%) | 1/7 (14.3%) | 2/11 (18.2%) | 0/7 | 0/5 | 0/12 | 2/23 (8.7%) |
| Clindamycin | 0/4 | 1/7 (14.3%) | 1/11 (9.1%) | 0/7 | 0/5 | 0/12 | 1/23 (4.4%) |

^2^ Ceftazidime and cefepime
